# Modeling robust COVID-19 intensive care unit occupancy thresholds for imposing mitigation to prevent exceeding capacities

**DOI:** 10.1101/2021.06.27.21259530

**Authors:** Manuela Runge, Reese A.K. Richardson, Patrick Clay, Arielle Eagan, Tobias M. Holden, Manisha Singam, Natsumi Tsuboyama, Philip Arevalo, Jane Fornoff, Sarah Patrick, Ngozi O. Ezike, Jaline Gerardin

## Abstract

In managing COVID-19 with non-pharmaceutical interventions, occupancy of intensive care units (ICU) is often used as an indicator to inform when to intensify mitigation and thus reduce SARS-CoV-2 transmission, strain on ICUs, and deaths. However, ICU occupancy thresholds at which action should be taken are often selected arbitrarily. We propose a quantitative approach using mathematical modeling to identify ICU occupancy thresholds at which mitigation should be triggered to avoid exceeding the ICU capacity available for COVID-19 patients. We used a stochastic compartmental model to simulate SARS-CoV-2 transmission and disease progression, including critical cases that would require intensive care. We calibrated the model for the United States city of Chicago using daily COVID-19 ICU and hospital census data between March and August 2020. We projected ICU occupancies from September to May 2021 under two possible levels of transmission increase. The effect of combined mitigation measures was modeled as a decrease in the transmission rate that took effect when projected ICU occupancy reached a specified threshold. We found that mitigation did not immediately eliminate the risk of exceeding ICU capacity. Delaying action by 7 days increased the probability of exceeding ICU capacity by 10-60% and this increase could not be counteracted by stronger mitigation. Even under modest transmission increase, a threshold occupancy no higher than 60% was required when mitigation reduced the reproductive number *R*_*t*_ to just below 1. At higher transmission increase, a threshold of at most 40% was required with mitigation that reduced *R*_*t*_ below 0.75 within the first two weeks after mitigation. Our analysis demonstrates a quantitative approach for the selection of ICU occupancy thresholds that considers parameter uncertainty and compares relevant mitigation and transmission scenarios. An appropriate threshold will depend on the location, number of ICU beds available for COVID-19, available mitigation options, feasible mitigation strengths, and tolerated durations of intensified mitigation.

## Introduction

In the first half of 2020, the global spread of SARS-CoV-2 left many countries with no option other than to shut down their economies and encourage people to isolate by staying home. In the United States (US), stay-at-home policies implemented in late March and April of 2020 reduced the number of new infections and deaths [1]. In mid-2020, US states began to relax their stay-at-home policies [1,2] despite a lack of effective treatments or a vaccine. In late 2020, many states experienced epidemic waves as large as, or larger than, their initial epidemics, putting renewed strain on hospital resources and requiring new mitigation measures [1–3].

Intensive care resources, particularly staffed beds and ventilators, are limited [4,5] especially in rural areas [6,7]. In early 2020, many intensive care units (ICUs) in the US and other countries operated near and above capacity limits [8–12]. To ensure continued life-saving care and a functioning health system, ICU occupancies must stay below capacity, and multiple guidelines for managing ICU capacities during COVID-19 surges have been formulated [5,13–15].

In response to fluctuations in SARS-CoV-2 transmission, states formulated COVID-19 response strategies to guide transitions between mitigation and relaxation policies [1]. These mitigation and relaxation policies defined setting-specific COVID-19 prevention measures such as occupancy limits for businesses, constraints on indoor activities, work from home recommendations, or population-wide stay-at-home orders (‘lockdowns’). For instance, in the US state of Illinois, thresholds used to spur increasing mitigation measures included test positivity rate (if surpassing 8%), increasing or decreasing trends in occupied hospital beds, and total ICU bed availability (if below 20%) [16]. The selection of robust yet sensitive thresholds to trigger a strategic mitigation response is challenging but critical, as health departments require time to appropriately prepare for and respond to a potential increase in transmission and hospital bed demand but prefer not to impose unnecessary mitigation (Fig 1). Thresholds that are too low could lead to premature restrictions or harmful effects on the economy and the community due to unnecessarily remaining under mitigation for too long. Thresholds that are too high could lead to late action, strained hospital resources, and elevated rates of severe COVID-19 cases and deaths.

**Fig 1.**
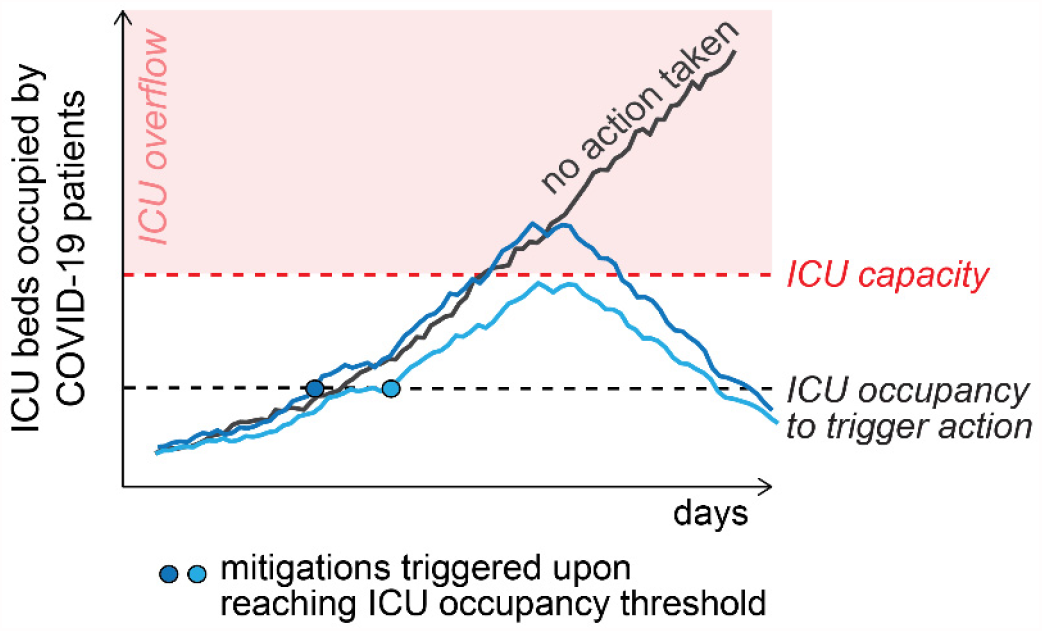
Conceptual visualization of ICU occupancy when mitigation is triggered compared to no action taken. The dashed red line indicates ICU capacity. The solid black line shows the scenario when no action is taken and ICU occupancy exceeds the ICU capacity, leading to ICU overflow (red area). The blue lines show two example scenarios in which mitigation was triggered at the same specified threshold (dashed black line). In one scenario (light blue) ICU overflow is prevented, and in the second scenario (dark blue) ICU capacity is still exceeded. This demonstrates that the effectiveness of mitigation measures is probabilistic, not absolute.

Thresholds for action (ICU occupancy levels that, when met, trigger more intense mitigation measures against transmission) should not be arbitrarily selected but rather be designed to meet COVID-19-related public health targets. Several modelling studies explored short-term forecasting of ICU occupancies [17–25] and mitigation strategies in relation to ICU capacities [17–19,26]. However, the criteria for selection of thresholds for action has not been assessed in greater detail.

This study investigates how ICU occupancy can be used as an indicator to drive mitigation decisions to avoid exceeding ICU capacity. We modeled COVID-19 transmission and disease progression under various levels of transmission increase, mitigation effectiveness, and mitigation timeliness, corresponding to the situation in Chicago, Illinois, in late 2020. The results of this analysis provide a quantitative approach for selecting robust thresholds in Chicago that can be applied in similar areas.

## Methods

### Study area

Chicago is an urban area of 2.7 million people in the US state of Illinois, and around 12% of the population is aged 65 years or older [27]. 17.4 inpatient medical/surgical (med/surg) hospital beds and 4.2 ICU beds are available per 10,000 population across 27 community hospitals [28]. After removing beds occupied by non-COVID-19 patients, the bed availability for COVID-19 patients is 6.7 med/surg beds and 2.2 ICU beds per 10,000 population.

The first SARS-CoV-2 infection was reported in Chicago in mid-January 2020 [21]. On March 21, 2020, a statewide stay-at-home order was announced to contain the spread of the virus. The stay-at-home order was gradually relaxed at the end of May 2020, and restaurants and recreational locations were allowed to reopen at the end of June [30]. Although the number of reported cases stayed relatively low during the summer, transmission increased during fall and on November 20, 2020, a second stay at home order was issued [31].

### COVID-19 transmission model

We used a stochastic Susceptible, Exposed, Infectious, and Recovered (SEIR) compartmental model, with additional compartments for the symptomatic subgroups (asymptomatic, pre-symptomatic, mild symptoms, severe symptoms), hospitalizations, critical cases that require treatment in ICUs, and deaths (Fig 2). Full model details are provided in the supplement (S1 Appendix). The model was implemented using the open source simulation engine Compartmental Modeling Software [32] combined with a simulation management framework in Python 3.9 [33] and post-processing in R 4.0 [34].

**Fig 2.**
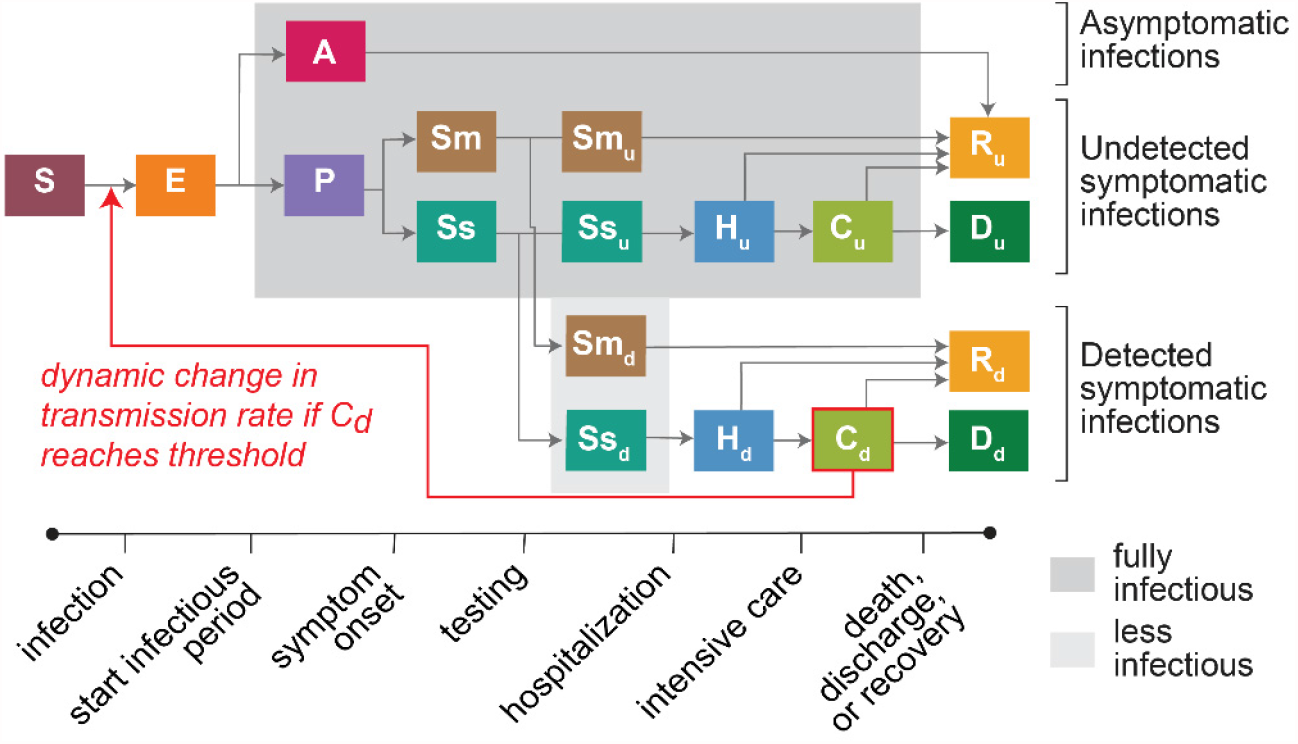
Structure of SARS-CoV-2 transmission and COVID-19 disease progression model. The compartments include Susceptible (S), Exposed (E), Asymptomatic (A), Pre-symptomatic (P), Mild symptomatic (Sm), Severe symptomatic (Ss), Hospitalized (H), Critical - intensive care (C), Deaths (D), Recovered (R). The subscripts d and u refer to infections detected and undetected by diagnostic testing, respectively. The red arrow shows the feedback loop from ICU COVID-19 occupancy within the C_d_ compartment (detected SARS-CoV-2 infections requiring intensive care) affecting the transmission rate that defines the transition from S to E.

Time-varying detection rates for severe and mild cases were taken from an analysis of Illinois case and death data [35] (S1 Fig 2-3). All Illinois data used for model parameterization, calibration, and validation were obtained from the Illinois Department of Public Health (IDPH). Time-varying fraction of hospitalized cases requiring critical care and time-varying case fatality rates were derived from the Illinois National Electronic Disease Surveillance System (I-NEDSS) [36] (S1 Fig 3-5). Lengths of stay in the hospital and ICU were informed by data from Northwestern Memorial Hospital in Chicago and from literature [37,38]. Other parameters were based on research studies outside Illinois (S1 Table 1).

Triggered mitigation measures after October 1, 2020, were applied as a decrease in transmission rate, non-specific to the mitigation measures that would cause this decrease (such as the closure of retail business, stricter mask-wearing protocols, or shelter-in-place). We set a feedback loop between the population in the critical detected (C_d_) compartment (COVID-19 ICU occupancy) and the transmission rate parameter, such that C_d_ triggers mitigation at specified occupancy thresholds (Fig 2).

### Model calibration and fitting to Chicago epidemic

Time-varying transmission rate prior to September 1, 2020, was fit to confirmed daily COVID-19 ICU census and COVID-19 med/surg hospital census in Chicago between February and August 2020 (Fig 3A, S1 Fig 6-8). The census data included all confirmed COVID-19 patients currently occupying ICU or med/surg beds in Chicago hospitals, and no data was available on location of residence for individual patients. Each of the two data series was smoothed with a 7-day centered moving average prior to comparison with simulation outputs. The time-varying reproductive number *R*_*t*_ was calculated from simulated daily new incident infections using the python module *epyestim* [39], which is based on the methodology from Cori et al. [40]. We specified a smoothing window of four weeks and kept the default serial interval and delay distribution as specified in [39]. The final *R*_*t*_ estimates were smoothed using a rolling average for 3 days to reduce high variation in these estimates (Fig 3).

**Fig 3.**
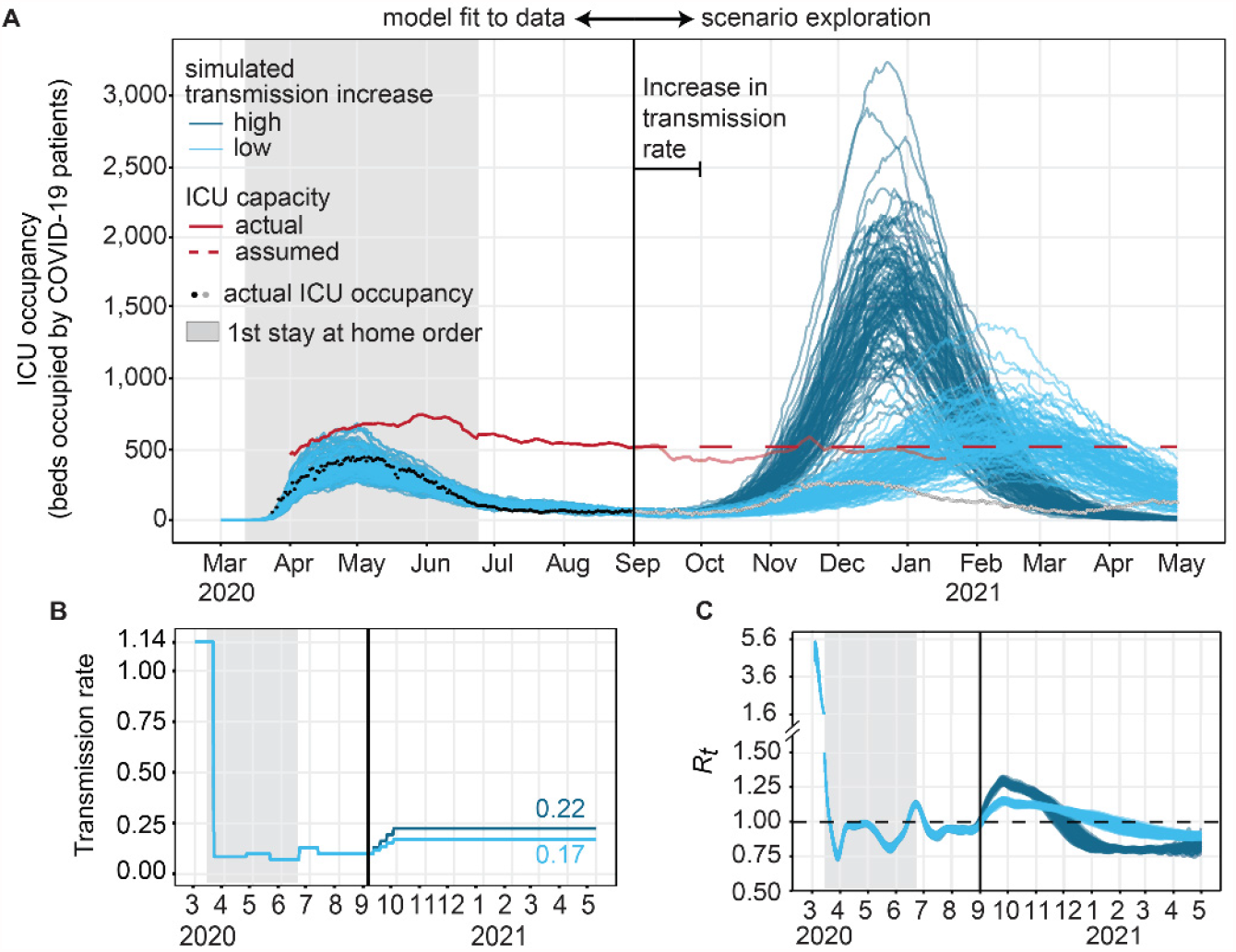
ICU occupancy over time, model fit, and increase in occupancy following an imposed increase in transmission. **A)** Predicted and observed daily ICU occupancy for Chicago in 2020. Black dots: actual COVID-19 ICU occupancy in Chicago. Red line: ICU bed capacity for COVID-19 patients, actual 7 day rolling average (solid) and model assumption (dashed) after September 2020. Blue lines: simulated trajectories under low (light blue) or high (dark blue) level of transmission increase and no triggered mitigation. The top 100 trajectories after fit to ICU census are shown (see S1 Fig 10 for full sample). **B)** Transmission rate parameter, which was fit to data before September 1, 2020, and gradually increased until October 1, 2020, to a higher (dark blue) or lower (light blue) target value. **C)** *R*_*t*_ estimated from simulated new infections.

In the fitting process, we first estimated the infection importation date (date with 10 infections), initial transmission rate, and transmission rate under mitigation for March 2020. We then fitted twice-monthly adjustments to the transmission rate between April and August. Other parameters were set to their mean value according to local data or epidemiological studies (S1 Table 2). Best fit parameter combinations were those that minimized the negative log likelihood of the simulated trajectories, based on a Poisson distribution. In the fitting, ICU census and med/surg census were weighted equally. The model fit was validated against COVID-19-like illness (CLI) hospital admissions data for Chicago hospitals and against COVID-19 deaths from I-NEDSS with Chicago listed as county or ZIP code of residence. To account for parameter uncertainty, we ran simulations with 400 unique parameter combinations using fitted parameter ranges and, for the data-informed parameters, values sampled from uniform distributions. We then chose 100 trajectories (unique set of parameters) that best fit the ICU census data and used these parameter sets in the later analysis. To describe the fitting accuracy we calculated the mean absolute error (MAE) [41], using the *metrics* R package [42], for the median prediction compared against the weekly moving average of the data (S1 Fig 7).

### Simulated scenarios

Based on daily ICU occupancies and total bed availability in Chicago, we calculated that in 2020, on average 44% of all ICU beds were occupied by non-COVID patients, theoretically leaving 56% of beds available for COVID-19 patients. The average number of ICU beds available for COVID-19 patients during the week immediately preceding September 15, 2020, (516 ICU beds) will be referred to as ICU capacity in this work. We assumed the capacity of 516 ICU beds to stay constant capacity during the simulation period, whereas in practice the capacity ranged between 407 to 744 beds on a seven-day rolling average (Fig 3A, S1 Fig 14).

We imposed a gradual increase in the transmission rate beginning on September 1, 2020, and leveling off on September 30 to allow reduction in transmission starting from October due to mitigation if occupancy thresholds were reached (Fig 3B). The levels of increase in transmission were selected such that, without any subsequent decrease in transmission, either half (‘low increase’) or all (‘high increase’) of the trajectories exceeded ICU capacity by January 1, 2021. This resulted in an increase in transmission by either 71% for the ‘low increase’ scenario and 126% increase for the ‘high increase’. The actual epidemic trajectory in Chicago between September and December 2020 was slightly above simulated trajectories under the lower level of transmission increase scenario (Fig 3A). For comparison, in an updated fitting iteration using data until December 2020, a 60% increase in the transmission rate was estimated between September and November for a slightly higher baseline transmission rate.

Between October 2020 and May 2021, mitigation (immediate reduction in transmission rate) was triggered either one or seven days after the COVID-19 ICU occupancy threshold was reached. Mitigation was simulated to reduce the transmission rate by 20, 40, 60, or 80% (‘weak’, ‘moderate’, ‘strong’, or ‘very strong’). Once applied, changes in transmission rate due to mitigation were never reversed. A table comparing the assumed transmission increase and reduction values to other studies is included in the Supplement (S1 Table 5).

We explored scenarios in which we varied the increase in transmission (two levels), mitigation effectiveness (four levels), and mitigation delay (two levels), and ICU occupancy threshold that triggered mitigation (eleven levels), resulting in a total of 176 unique scenarios (Table 1). Each scenario was simulated with 400 sets of sampled parameters, drawn from uniform distributions (S1 Table 6). The top 100 trajectories that best fit to ICU census data up to September 1, 2020, were retained for each of the 176 scenarios. Trajectories in which the ICU occupancy threshold to trigger mitigation was not reached by May 2021 were excluded (5-6%, S1 Table 6). The sampled parameters were summarized using the mean and 90% prediction interval (PI).

**Table 1.**
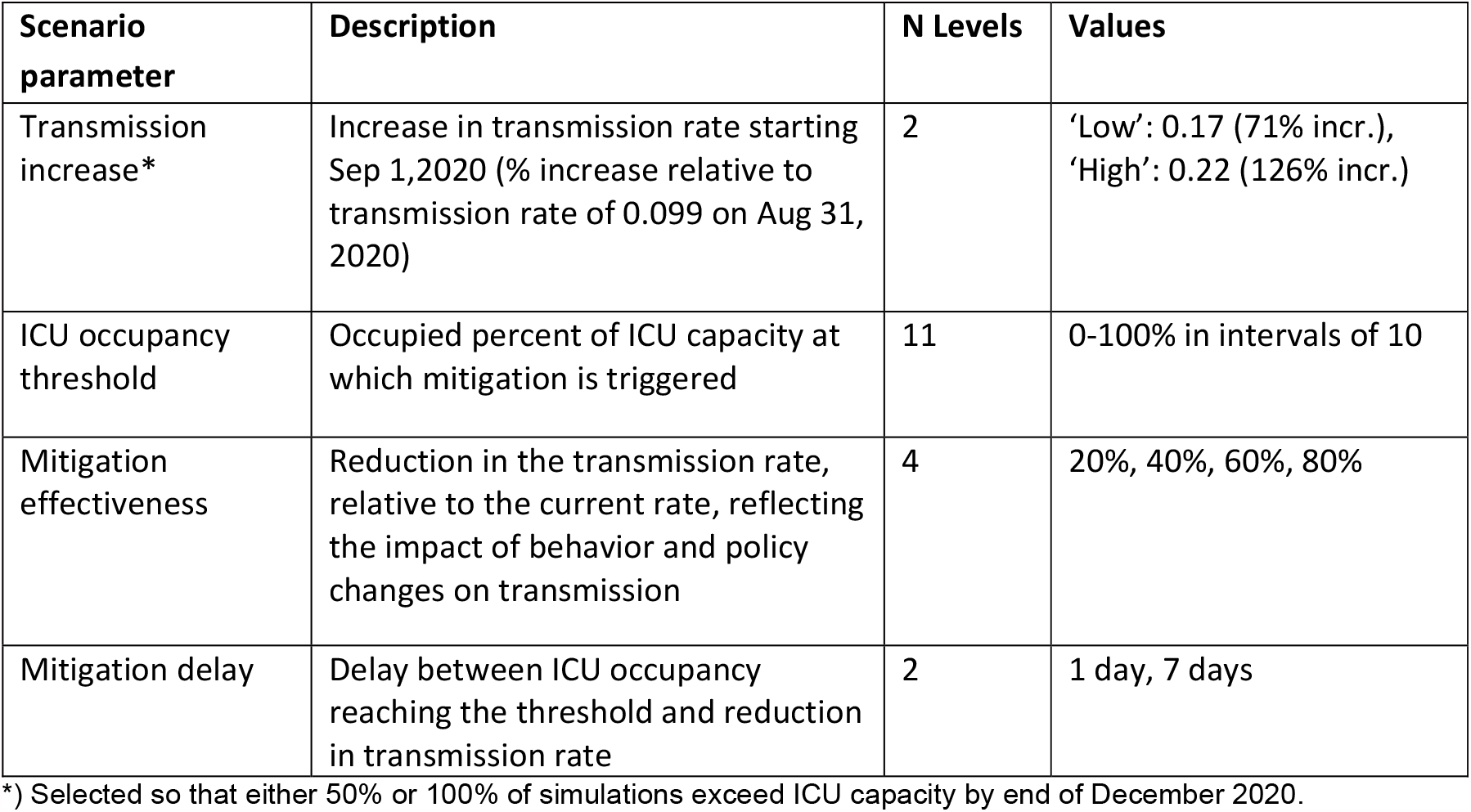
Overview of simulated scenarios

Daily COVID-19 ICU occupancy was the primary outcome in the analysis. The probability of exceeding ICU capacity (for COVID-19 patients) was calculated by dividing the number of trajectories that exceeded COVID-19 ICU capacity by the total number of trajectories that reached the COVID-19 ICU occupancy threshold to trigger mitigation. To account for different numbers of trajectories reaching the ICU trigger (S1 Fig 11, S1 Table 6), 50 trajectories were resampled 50 times allowing for replacements.

## Results

### Simulating a September 2020 epidemic wave in Chicago

We fit a compartmental model of SARS-CoV-2 transmission (Fig 2) to hospitalization and intensive care unit census data from Chicago between March and August 2020 (Fig 3A, S1 Fig 6-7). The fitted infection importation date was February 28, 2020, with an initial transmission rate of 1.14 and reproductive number *R*_*0*_ of 5.00 (90% prediction interval (PI) 4.57-5.41). After the stay-at-home order starting on March 22, 2020, we estimated a 92.5% reduction in the transmission rate (Fig 3B), reducing the time-varying reproductive number (*R*_*t*_) to 0.74 (90% PI 0.72-0.77) (Fig 3C). At the end of the fitted period on September 1, 2020, the transmission rate was estimated at 0.099 and the reproductive number at just around one (*R*_*t*_ = 0.99, 90% PI 0.96-1.03). The model captured trends in ICU occupancies reasonably well with an average MAE of 44 ICU beds occupied (range across trajectories 27-66) between March to August 2020.

To test the success of using ICU occupancy to trigger mitigations, we implemented two levels of an increase in transmission rate in September 2020. Without mitigation, the transmission increase led to a *R*_*t*_ of 1.14 (90% PI 1.12 to 1.16) for the lower level of increase and a *R*_*t*_ of 1.28 (90% PI 1.25 to 1.30) for the higher level of transmission increase on October 1, 2020. ICU occupancy increased until peaking in mid-December at the earliest and mid-February 2021 at the latest, depending on the level of transmission increase and parameter sample. The mean peak ICU occupancy reached 663 beds (90% PI 421-948) at the lower transmission increase and 1656 beds (90% PI 1096-2236) at the higher transmission increase, compared with ICU capacity of 516 beds. Across both levels of transmission increase, the projected peak ICU demand was 1.2-3.2 times more ICU beds needed than available.

### Preventing ICU overflow strongly depends on ICU occupancy threshold for action

Mitigation was triggered when the simulated ICU occupancy reached a pre-defined threshold relative to the ICU capacity (Fig 4). Stronger mitigation (>60% reduction in transmission rate) led to lower peak ICU occupancy and peak occurred sooner (Fig 4A, 4B). At the higher level of transmission increase, new infections dropped after mitigation was triggered (S1 Fig 13) and estimated *R*_*t*_ reached a minimum after around two weeks, before increasing again and leveling off below 1. The estimated *R*_*t*_ varied slightly across the simulated scenarios, and at the high transmission increase scenario, the *R*_*t*_ two weeks prior mitigation was estimated at of 1.23 (90% PI: 1.18-1.29) and was reduced to 1.03 (90% PI: 0.99-1.07) at weak, to 0.92 (90% PI: 0.88-0.96) at moderate, to 0.75 (90% PI: 0.71-0.79) at strong, and to 0.47 (90% PI: 0.40-0.54) at very strong mitigations (Fig 4D, S1 Fig 18).

**Fig 4.**
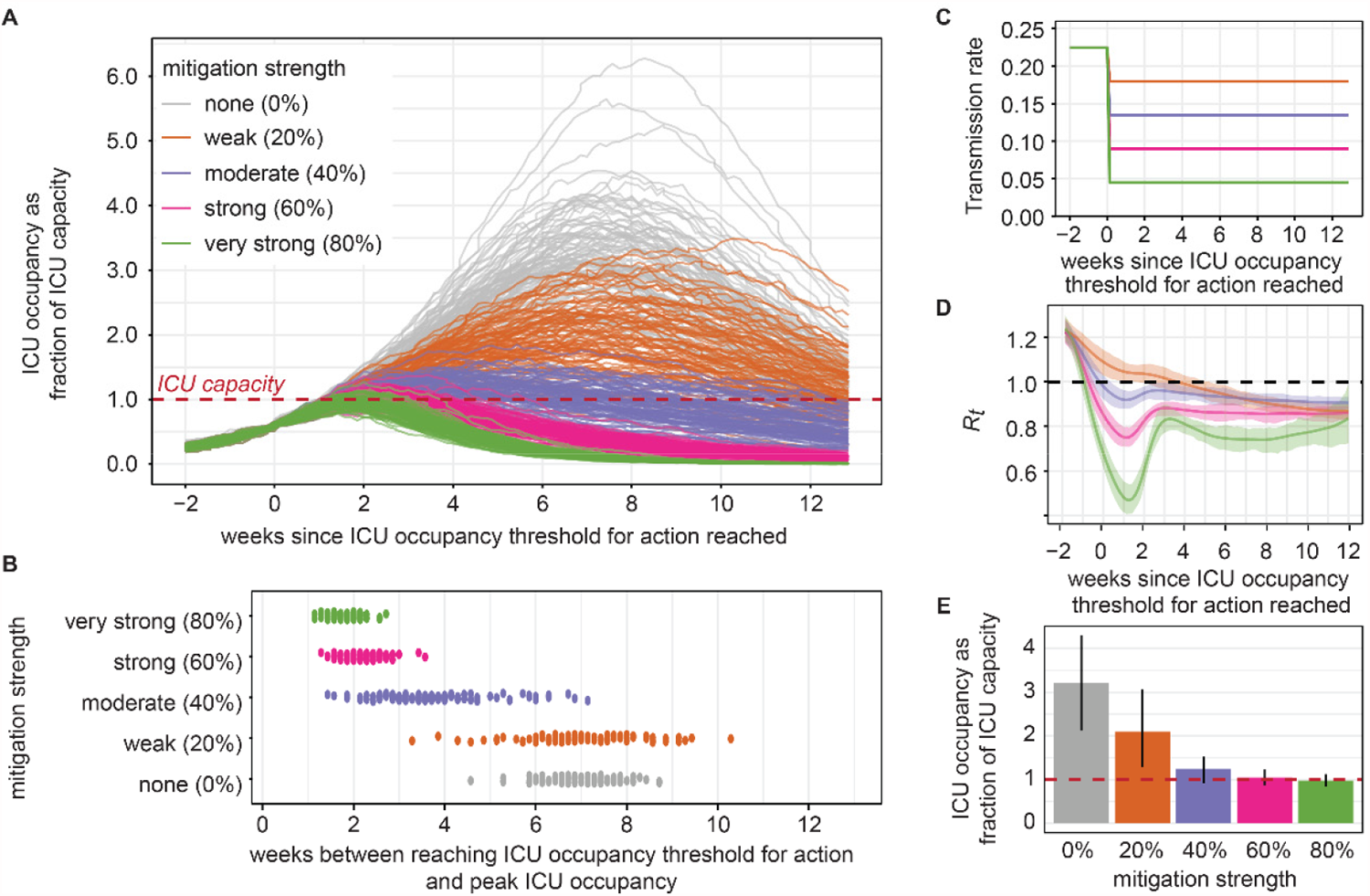
Projected outcomes under no mitigation compared to four mitigation strengths. **A)** Projected ICU census over time relative to the time when ICU occupancy reached the ICU occupancy threshold. **B)** Timing of peak ICU occupancy relative to the time when ICU occupancy reached the ICU occupancy threshold. **C)** Reduction in transmission rate due to immediate mitigation after reaching a 60% occupancy threshold. **D)** Estimated mean *R*_*t*_ with 90% PI uncertainty intervals. Note, maximum *R*_*t*_ might occur more than 2 weeks prior to mitigation depending on timing of triggered mitigation. **E)** Peak ICU occupancy, with mean and 90% PI error bars. All mitigations were implemented 1 day after reaching a 60% ICU occupancy threshold at varying mitigation strength (% reduction in transmission). The figure shows the scenario at higher transmission increase and the version for lower transmission increase shown in S1 Fig 12.

Compared to no mitigation, immediate mitigation decreased peak ICU occupancy by 34.5% (90% PI: 11.8-52.4%) under weak mitigation, by 59.7% (90% PI: 43.6-71.4%) under moderate mitigation, by 65.6% (90% PI: 50.3-75.2%) under strong mitigation, and 68.5% (90% PI: 54.4-77.4%) under very strong mitigation (Fig 4E).

ICU occupancy continued to grow for a short time after mitigation was imposed (Fig 5A). At the same mitigation strength, peak ICU occupancy was reached at a similar length of time (12 days) after mitigation regardless of the threshold ICU occupancy. Lower occupancy thresholds for triggering mitigation led to a lower probability of ICU overflow and lower peak ICU occupancy (Fig 5B).

**Fig 5.**
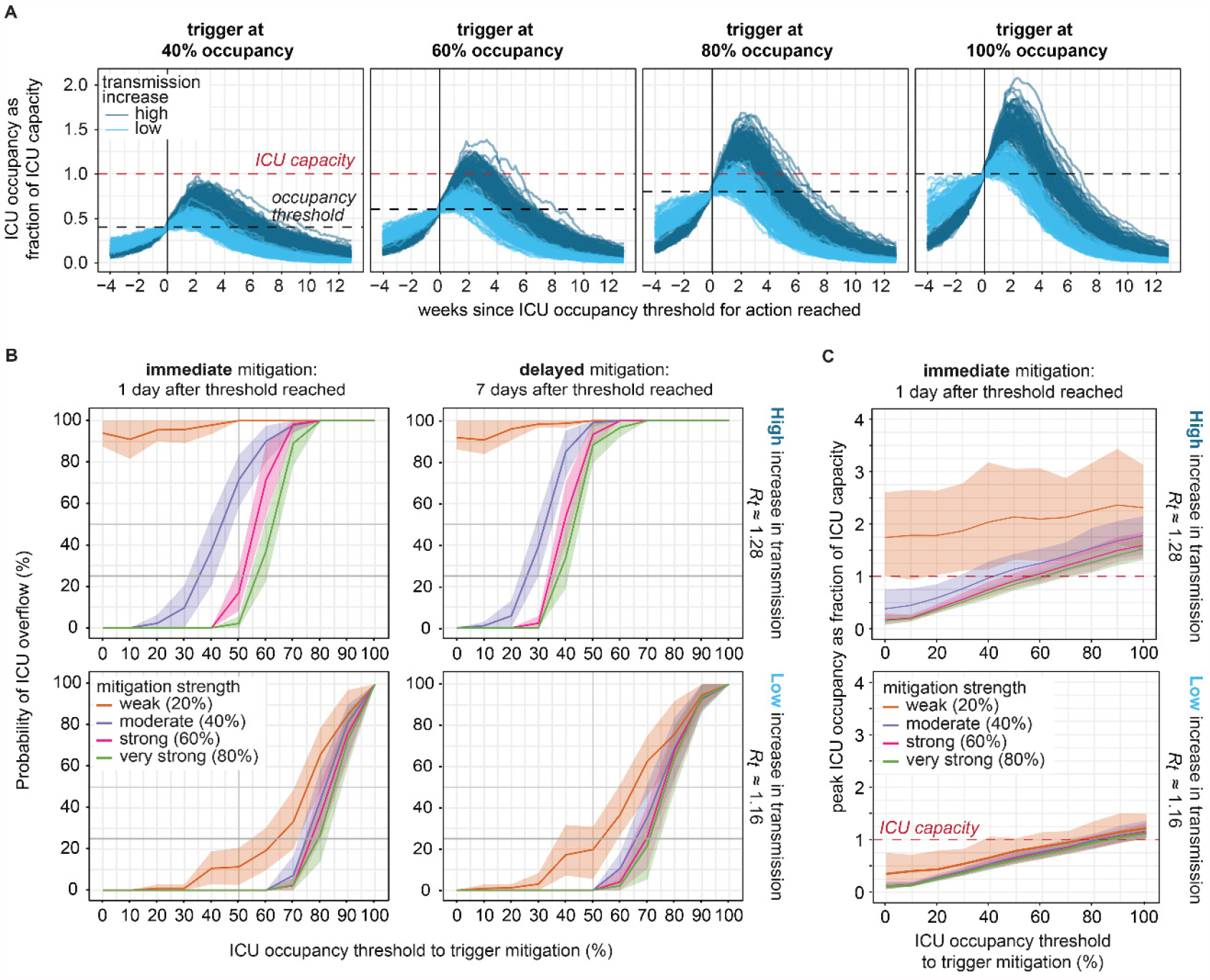
Probability of exceeding ICU capacity by ICU occupancy threshold, mitigation strength and timing. **A)** Projected ICU occupancy over time under various ICU occupancy thresholds to trigger mitigation. All scenarios show effects of immediate strong mitigation (60% reduction in transmission rate) and include trajectories simulated under either low (light blue) or high (dark blue) levels of transmission increase. **B)** Probability of ICU overflow under higher (top) or lower (bottom) level of transmission increase and immediate (left) mitigation or delayed (right). The lines and shaded areas show the mean, minimum and maximum probability obtained from resampling 50 trajectories 50 times. **C)** Peak ICU occupancy by ICU occupancy threshold, mitigation strength, and level of transmission increase, for scenarios of immediate mitigation.

We calculated the probability of exceeding ICU capacity under different possible ICU occupancy thresholds at which mitigation was dynamically triggered (Fig 5B). We compared the probabilities by transmission level, mitigation strengths, and delay between trigger and reduction in transmission due to mitigation. The probability of overflow increased with a higher ICU occupancy threshold. The probability was, on average across the ICU occupancy thresholds, 33% (range across mitigation strengths: 17%-63%) higher at the higher level of transmission increase compared to the lower level. Weak mitigation (20% decrease in transmission rate), where *R*_*t*_ was not reduced below 1, had a substantially higher probability of overflow than the other mitigation levels, and differences were greater for the higher level of transmission increase.

At the lower level of transmission increase, the probability of ICU overflow was almost identical for moderate, strong, and very strong mitigation, whereas at the higher transmission increase the difference between the mitigation levels was more pronounced with consistently high probability of overflow for weak mitigation. At the lower level of transmission increase, the probability of ICU overflow increased at thresholds above 30% occupancy when mitigation was weak, whereas when mitigation was moderate or stronger, the probability remained near zero until an ICU occupancy threshold of 60-70% after which the probability increased sharply. The incremental difference in the overflow probability between weak and moderate mitigation was 10% and less than 3% for the other mitigation strengths at the low increase level. In comparison, at the high increase level, the probability of exceeding capacity increased at thresholds above 40-50% occupancy and reached 100% for occupancy thresholds above 60-70% at strong and very strong mitigation. The difference in the overflow probability was 42% between weak and moderate mitigation, 16% between moderate and very strong mitigation, and negligible between strong to very strong mitigation (<1%) (Fig 5, Fig S1 Fig 14).

A delay of seven days shifted the probability curves to the left, with higher probability of overflow at each of the ICU occupancy thresholds. For instance, at an 80% ICU occupancy threshold and very strong mitigation, the probability of overflow increased from 41.8% to 89.6% under the lower level of transmission increase when mitigation was delayed by seven days. At the higher level of transmission increase, a 60% occupancy threshold and very strong mitigation had 37% probability of overflow if mitigation was immediate but 97.4% probability of overflow if mitigation was delayed. When assessing mitigation strengths against delay, the probability of overflow was higher for strong mitigation that was delayed by seven days compared to moderate mitigation with immediate action. For a hypothetical risk tolerance of 25% probability of ICU overflow, the required ICU occupancy thresholds for action were 40 to 60% across the tested scenarios.

A policy of 80% ICU occupancy to trigger mitigation did not prevent exceeding capacity, as mean peak ICU occupancy was at or above ICU capacity for all mitigation strengths and both levels of transmission increase. A 60% ICU occupancy threshold for mitigation was barely sufficient for preventing ICU overflow: mean peak ICU occupancy remained below capacity for the lower transmission increase at all mitigation strengths but remained below capacity for the higher transmission increase only under very strong mitigation. Under the higher level of transmission increase, weak mitigation (20% reduction in transmission) could not contain mean peak ICU occupancy to below ICU capacity regardless of the ICU occupancy that triggered mitigation (Figure 5C). Above mitigation strength of 40%, stronger mitigation did not substantially reduce peak ICU occupancy at either level of transmission increase.

### Mitigation strength and occupancy threshold determine time spent above capacity

For simulation trajectories where ICU capacity was exceeded, we measured the number of days in each trajectory where ICU occupancy exceeded capacity (Fig 6, S1 Fig 14). Without mitigation, the duration above ICU capacity was on average 81 days at the higher level of transmission increase and 68 days at lower transmission increase. Under immediate mitigation, the average duration above ICU capacity for the higher transmission increase was reduced to 78 days with weak mitigation and further decreased under stronger mitigation (to 43, 21, 14 days for moderate, strong, and very strong), averaged across ICU occupancy thresholds. The corresponding average duration above capacity for the lower transmission increase were 26, 14, 10 and 8 days for weak, moderate, strong and very strong mitigation respectively. Higher ICU thresholds, and hence later action, resulted in longer duration above capacity. A delay of seven days in mitigation did not substantially extend the time above capacity beyond the seven days among those trajectories that exceeded the capacity (Fig 6B).

**Fig 6.**
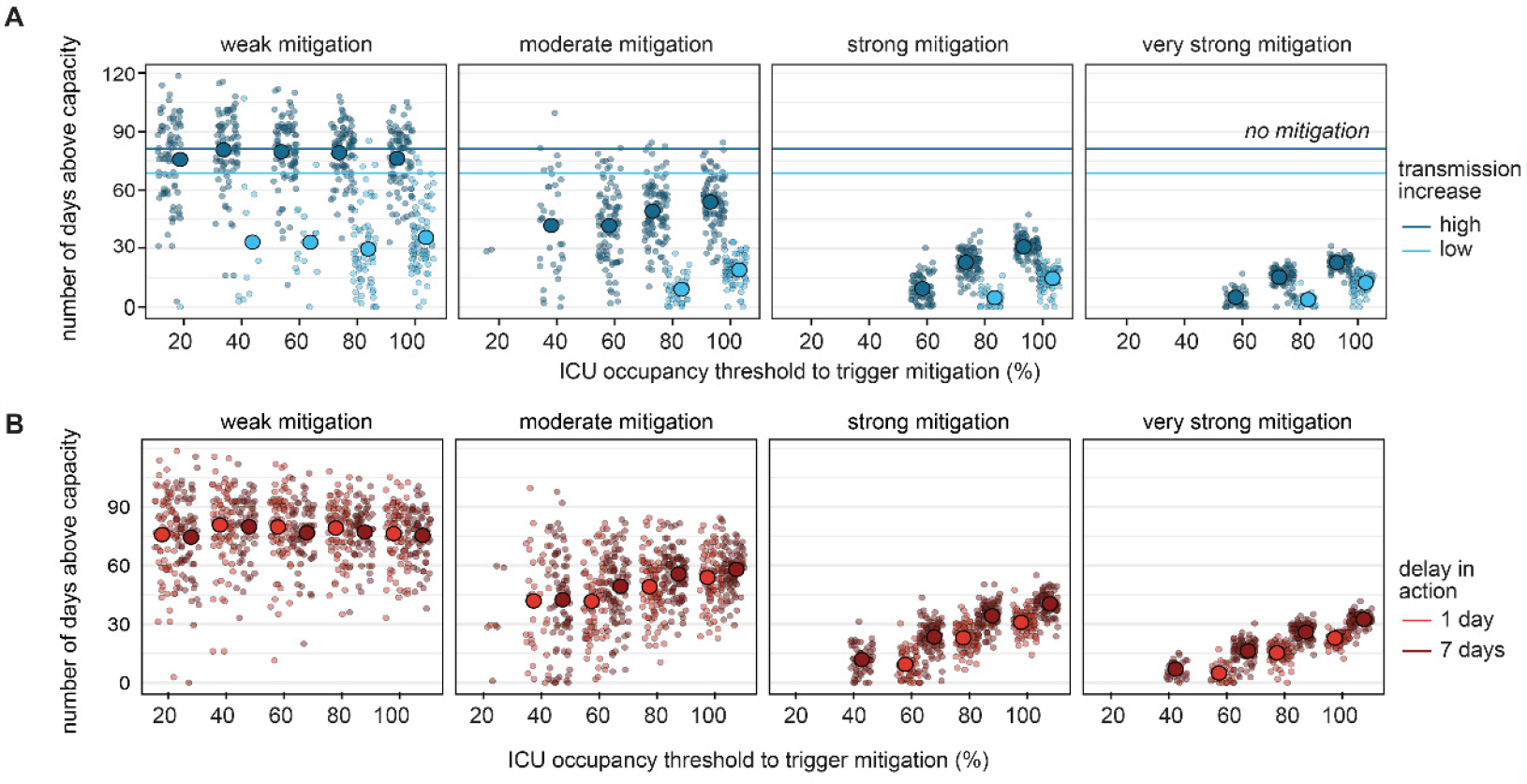
Number of days spent above ICU capacity under various possible response scenarios. **A)** Number of days above ICU capacity by transmission increase level with immediate mitigation. **B)** Number of days above ICU capacity by mitigation timeliness under high transmission increase. Shown: single trajectories and aggregated mean. The number of trajectories included in each scenario group is shown in S1 Fig 15.

## Discussion

We developed a model of SARS-CoV-2 transmission in Chicago to explore how ICU occupancy can be used as an indicator for triggering new mitigations in response to increasing transmission. ICU occupancy is a late indicator for SARS-CoV-2 transmission since ICU admission lags symptom onset by 10 days [37], which lags infection by up to 14 days [43]. However, we find that ICU occupancy can still be a critical guide for policy action if action is taken promptly, mitigation reduces *R*_*t*_ to below 1, and occupancy thresholds for action are conservatively low.

In an initially expanding epidemic, ICU occupancy of COVID-19 patients will continue to increase for around two weeks after imposing mitigations. Higher ICU occupancy thresholds for action thus increase the probability of overshooting ICU capacity during those two weeks. Furthermore, mitigation measures could be delayed to give individuals and businesses warning in advance of changing policies. Scaling up of hospital beds and staff might require even longer notice times of three to four weeks [44,45]. These delays would result in additional hospitalizations and increase the probability of ICU overflow. We found that mitigation strength could not compensate for a delay in action. Other researchers have also noted a critical window during which policies need to be implemented and have shown that even short delays can result in substantial increase in infections [46]. If immediate action (i.e. less than 3 days) is not feasible, an alternative would be to reduce the threshold for triggering mitigation to allow more time for planning and implementation. Anticipating a delay is crucial when selecting a threshold if ICU capacity is not to be exceeded.

This study models mitigation as an abstracted decrease in transmission rate. In current practice, mitigation is achieved through a mix of social distancing, masking, diagnostic testing, isolation, and contact tracing [47]. The strongest mitigation considered in this study reduced transmission to levels below what was observed in Chicago during the stay-at-home order in March 2020, when mitigation relied heavily on social-behavioral changes as access to diagnostic testing was limited and contact tracing had yet to be implemented. Practical implementation of the “very strong” mitigation modeled in this study would therefore require both interventions that reduce contact rates and interventions to promote early diagnosis and isolation. Studies in the US and Canada estimated that social distancing alone would be not enough to prevent ICU overflow during the first epidemic wave of 2020 [19,20]. Higher mitigation strength might be easier to achieve in higher populated areas than in more sparsely populated areas with less mobility and already relatively low contact rates. However, determining an expected reduction in transmission given specific mitigation plans is challenging since transmission is influenced by many behavioral factors that vary geographically, demographically, and over time. Reductions in between 20% and 95% have been estimated across a range of studies (S1 Table 5).

In Chicago and other regions in Illinois, the ICU occupancy threshold to spur transition to the next COVID-19 mitigation phase was at 80% of total occupancy (20% total availability), corresponding to around 40% occupancy of beds available for COVID-19 patients [16]. In theory, different geographical areas could have different ICU occupancy thresholds for action tailored to their specific context, determined by factors such as current *R*_*t*_, anticipated population behavior, ICU flexing capacity, and overall risk tolerance for exceeding ICU capacities. In practice, using region-specific thresholds risks an uncoordinated response [48], and mitigation might not be as effective due to spillover effects across neighboring regions. During Chicago’s October 2020 epidemic wave, mitigation was implemented on November 20 [31] when the COVID-19 ICU occupancy was 53% (S1 Fig 16). However, *R*_*t*_ had already begin to decrease prior to implementation of official mitigation measures as individual action preceded government policy (S1 Fig 17).

Our suggested threshold of 60% for Chicago aligns with thresholds used in a modeling study that simulated multiple on-off cycles based on a fixed 50% ICU occupancy threshold [26]. Another modeling study evaluated an ICU threshold system that defined 30% occupancy as moderate risk, 30-60% occupancy as higher risk, and above 60% as very high risk [18]. This study found that ICU-based thresholds would be overly restrictive, suggested a strategy based on COVID-19 hospital admissions [18], and chose 80 hospital admissions per day as a more appropriate trigger [17]. The Illinois Department of Public Health defined an 80% threshold on total ICU occupancy, translating to a occupancy threshold of around 50% for COVID-19 patients when assuming a maximum 60% occupancy by non-COVID-19 patients. In our analysis, a 40% occupancy threshold is associated with relative low probabilities of overflow. In practice, local health departments monitor multiple indicators and the decision to act depends on the combination of all indicators or the most restrictive one at a given time. Unfortunately, all these measures (case counts, case rates, test positivity rates, hospital admissions, hospital census, ICU census, and deaths) are limited in providing timely and accurate trends as they are biased, noisy, or lag infection by several weeks.

We defined ICU capacity as the number of staffed, supplied beds available to treat critically ill COVID-19 patients, and we assumed COVID-19 ICU capacity (difference between total capacity and non-COVID-19 occupancy) stayed constant. Historical trends, however, showed fluctuations in ICU capacity, reflecting both non-COVID-19 use and hospitals following COVID-19 ICU response strategies to scale up or ramp down beds in response to trends [5,14,15]. For instance, one strategy stretches ICU capacity 20% above normal by using existing staff and resources to respond to minor surges in ICUs [5]. Rescheduling elective surgeries also impacts bed availability and can increase the number of beds available for COVID-19 patients [49]. Our results therefore could overestimate the probability of exceeding capacity if there is flexibility to increase capacity when occupancy is high. However, flexing ICU capacity puts additional strain on the health care system and may potentially impact the quality of care. Conversely, our analysis would underestimate the probability of exceeding ICU capacity if non-COVID-19 related admissions increased simultaneously with SARS-CoV-2 transmission.

At the state level, COVID-19 ICU management also includes patient transfers across hospitals and regions. Transfers are more common from smaller hospitals in more rural areas to larger specialized hospitals in urban areas than the reverse [49]. In urban areas, high ICU occupancies could therefore contain a substantial contribution of patients residing in the surrounding region, and near-capacity or overflow of urban ICUs would put additional strain elsewhere. At Northwestern Memorial Hospital, one of the largest hospitals in Chicago, 25-30% of COVID-19 admissions were not Chicago residents (S1 Fig 18). Hence, while capacities are higher in urban areas, accounting for potential patient transfers may require lowering the threshold for action. As occupancy nears capacity, and patients are admitted to nontraditional ICU areas or transferred to hospitals outside the region [49], ICU data becomes less reliable as an epidemic indicator for local transmission.

The model does not include vaccination, and we assume 34% of hospitalized COVID-19 patients will require ICU care [50]. As vaccine programs are scaled up, hospital and ICU admissions will substantially decrease and the demographics of admitted patients may also shift. Younger patients may reside longer in the ICU than the elderly if elderly patients are more likely to move to hospice care or are less likely to survive COVID-19. The model also did not include shifting virulence due to spread of new SARS-CoV-2 variants. Vaccination, changing demographics, and changing virulence may mean that ICU occupancy is no longer a good indicator of transmission and hence should not be used to make mitigation decisions. Whether and when to implement mitigation will need to increasingly depend on more direct measures of transmission. Nevertheless, it remains crucial to monitor COVID-19 hospitalizations and ICU occupancies and to have fail-safe thresholds in place to allow timely action to prevent severe illness and deaths.

## Conclusions

We used a SARS-CoV-2 and COVID-19 disease transmission model to evaluate how ICU occupancy can be used as an indicator for triggering new mitigations in response to increasing transmission in Chicago, USA. The model suggests that a threshold of at most 60% ICU occupancy can reliably prevent exceeding capacity, and amount of transmission increase, mitigation strength, and anticipated delays in mitigation effects are important factors when selecting thresholds. In each area, the appropriate threshold will further depend on the options available for mitigation, feasible mitigation compliance levels, and tolerable durations of intensified mitigation.

## Data Availability

The simulation and analysis code are publicly available on GitHub under https://github.com/numalariamodeling/ICUtrigger_covid_chicago_paper_2021. The simulation outputs are stored on Zenodo with DOI:10.5281/zenodo.5018779. The COVID-19 transmission model is maintained under https://github.com/numalariamodeling/covid-chicago. Public data is available on the IDPH website (https://dph.illinois.gov/covid19). Illinois hospital census data and I-NEDSS data can be received from IDPH upon reasonable request. All other data are publicly available as mentioned in the text.

https://doi.org/10.5281/zenodo.5018779

## Abbreviations

CMS: Compartmental Modeling Software
ICU: Intensive care unit
IDPH: Illinois Department for Public Health
I-NEDSS: Illinois National Electronic Disease Surveillance System
Med/surg: Medical/surgery hospital inpatient beds
PI: Prediction interval
SARS-CoV-2: Severe acute respiratory syndrome coronavirus 2
*R*_*t*_: Time varying reproductive number (also effective reproductive number *R*_*eff*_)

## Author Contributions

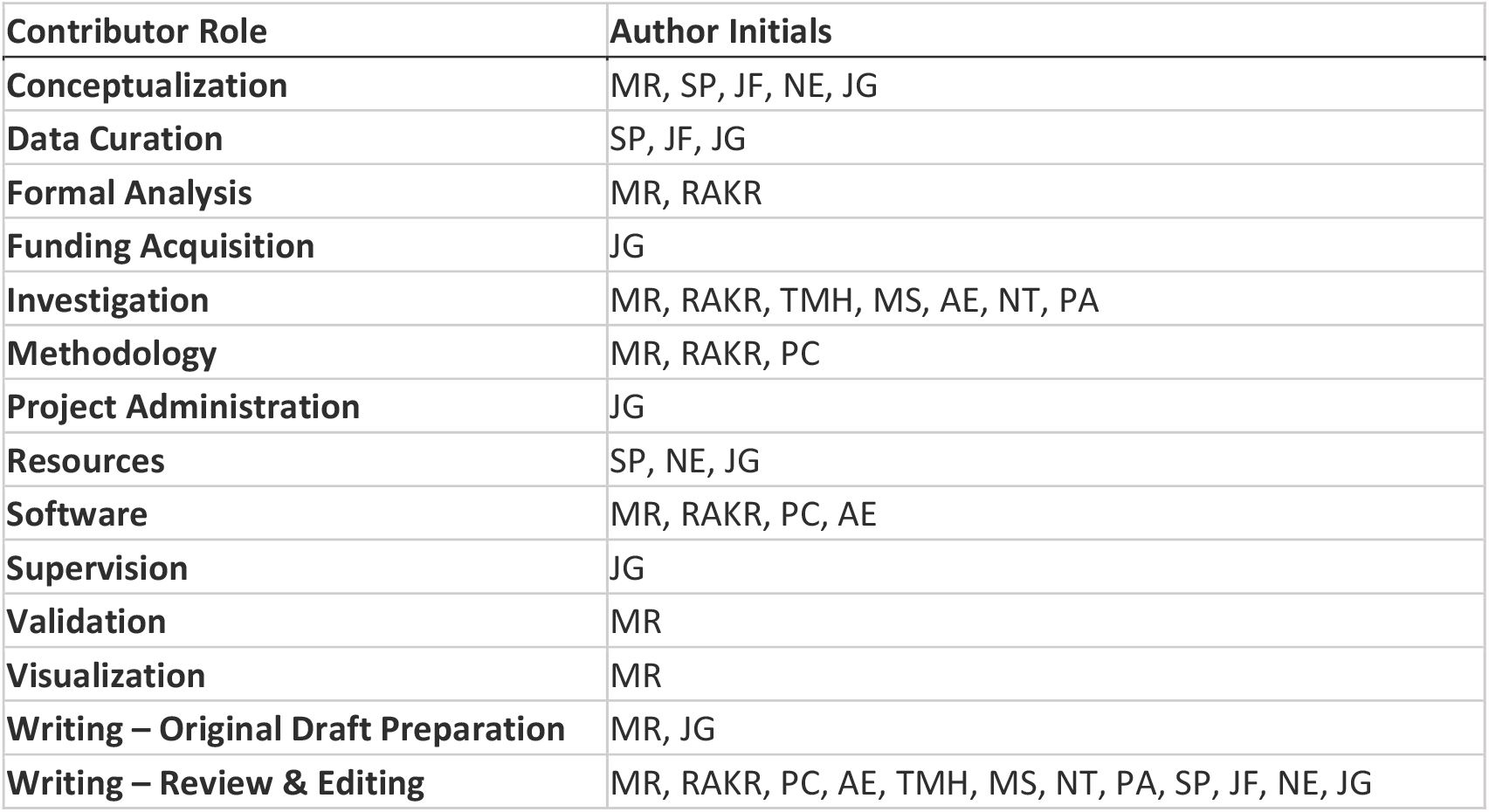

## Acknowledgements

We thank Chinyere Alu, Stacey Hoferka Jensen, Megan Patel, and Dejan Jovanov from IDPH and Michele Atkinson and Sara Rogers from Civis Analytics for data extraction and data management. We thank Stephen Hoover, Arthi Ramachandran, and Jackson Lee of Civis Analytics for technical assistance in developing the simulation software. We thank Ross York-Erwin from Northwestern Memorial Hospital for data insights and helpful discussions. We thank Janna Nugent and Scott Coughlin from Northwestern Research Computing for building a CMS docker on Linux and technical support on HPC. We thank Chris Lorton of the Institute for Disease Modeling for assistance on using the CMS software. We thank Ron Ackermann, Abel Kho, Nicholas Soulakis, and Theresa Walunas of Northwestern University for facilitating the connection with IDPH. We thank the Illinois COVID-19 Task Force: Sarah Cobey, Crystal Son, Jonathan Ozik, Charles Macal, Sergei Maslov, and Nigel Goldenfield for helpful discussions as well as Katie Gostic, Frank Wen, Sylvia Ranjeva, Lindsay Keegan, and Wayne Duffus. We thank Ariel Chandler, Brittany Hagedorn, Farhad Ghamsari, Aadrita Nandi, Ifeoma Ozodiegwu, Kamaldeen Okuneye, Keith Walewski, Kevin Yu, Kristyn Krolikowski, Tracy Guo, Zara Saldanha, and Garrett Eickelberg for technical and management support during the pandemic early in 2020. We thank Sebastian Rodriguez and Ben Toh for helpful comments on the draft manuscript.

This research was supported in part through the computational resources and staff contributions provided for the Quest high performance computing facility at Northwestern University, which is jointly supported by the Office of the Provost, the Office for Research, and Northwestern University Information Technology.

## Data availability

The simulation and analysis code are publicly available on GitHub under https://github.com/numalariamodeling/ICUtrigger_covid_chicago_paper_2021. The simulation outputs are stored on Zenodo with DOI:10.5281/zenodo.5018779. The COVID-19 transmission model is maintained under https://github.com/numalariamodeling/covid-chicago.

Public data is available on the IDPH website (https://dph.illinois.gov/covid19). Illinois hospital census data and I-NEDSS data can be received from IDPH upon reasonable request.

All other data are publicly available as mentioned in the text.

## Supplementary files

S1 Appendix: Technical supplement and additional results figures.

