## Supplementary material for "Modeling robust COVID-19 intensive care unit occupancy thresholds for imposing mitigation to prevent exceeding capacities": S1 Appendix

### Model details

The susceptible population is infected at a constant rate determined by the transmission probability and contact rate between the susceptible and infectious population. After a latent period of 3-4 days [1], the exposed population becomes infectious either asymptomatic or pre-symptomatic and after another 3-4 days, pre-symptomatic cases develop mild or severe symptoms [2]. Mild symptomatic cases recover at the same rate as asymptomatic cases with a recovery of around 10 days. Severe symptomatic cases are hospitalized after 3-6 days [3] and after 4-6 days either recover [3] or develop critical illness (ICU patients) [4]. Critical patients recover after 8-10 days [5] or die after 4-6 days [6]. The infectious population included all asymptomatic, pre-symptomatic, mild symptomatic, severe symptomatic infections, as well as hospitalized and critical cases that are undetected. We assumed a 70% to 100% reduction in infectiousness for the detected infectious population to account for isolation, whereas detected hospitalized and critical cases do not transmit. We assumed full immunity for recovered infections during the simulation timeframe. Vaccinations and variants of concern were not included (S1 Fig 1). A description of the transition rate parameters is shown in S1 Table 1 and a description of the model parameters and sources, either obtained from clinical or epidemiological studies or by Illinois specific data is shown in S1 Table 2.

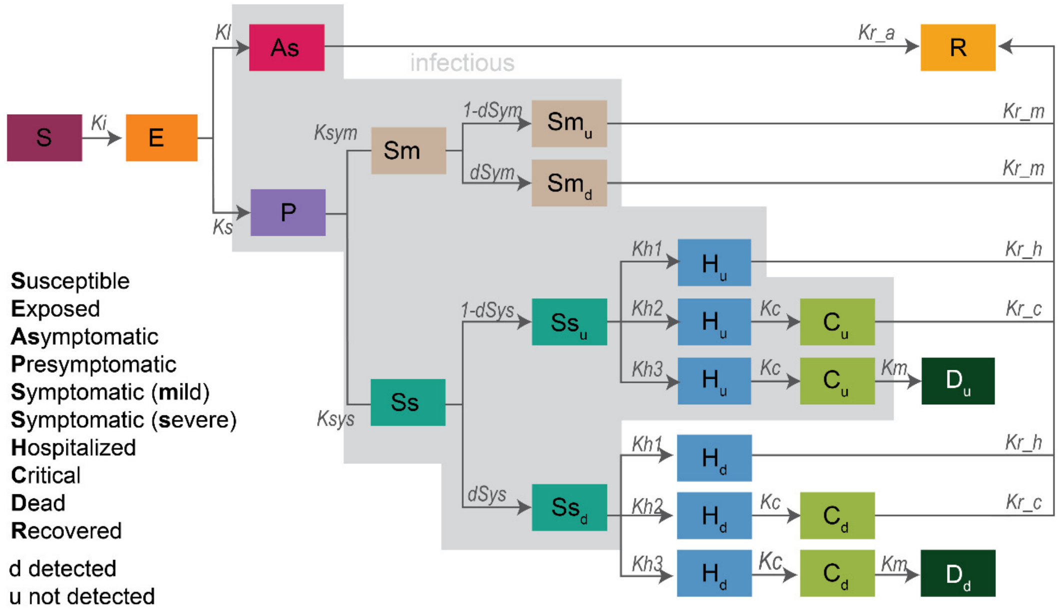

**S1 Fig 1. Full model structure and parameters.**

The grey shaded compartments are infectious, whereas compartments for detected populations have reduced infectiousness ($Sm_{det}$ and $Ss_{det}$) and infected hospitalized population was assumed to not transmit to the general population when detected. The model was implemented in the compartmental modeling software [7] and the python simulation framework is public available under https://github.com/numalariamodeling/covid-chicago.

**S1 Table 1.** Model transitions

| Parameter | Description | Transition rate |
|---|---|---|
| $Ki$ | Transmission rate | / |
| $Kl$ | Progression to asymptomatic | $\frac{1-f_S}{t_{AsP}}$ |
| $Ks$ | Progression to pre-symptomatic | $f_S$ / $t_{AsP}$ |
| $Ksym$ | Progression to mild symptomatic | $f_{Sm}$ / $t_{AsP}$ |
| $Ksys$ | Progression to severe symptomatic | $f_{Ss}$ / $t_{AsP}$ |
| $dSym$ | Detection of mild symptomatic | $d_{Sm_t}$ / $t_{Smdet}$ |
| $dSys$ | Detection of severe symptomatic | $d_{Ss_t}$ / $t_{Ssdet}$ |
| $Kh1$ | Hospitalizations of severe symptomatic that recover | $f_{H_t}$ / $t_H$ |
| $Kh2$ | Hospitalizations of severe symptomatic that progress to critical | $f_{C_t}$ / $t_H$ |
| $Kh3$ | Hospitalizations of severe symptomatic that die | $f_{D_t}$ / $t_H$ |
| $Kc$ | Progression to critical | 1 / $t_C$ |
| $Km$ | Death | 1 / $t_D$ |
| $Kr\_a$ | Recovery rate asymptomatic | 1 / $\gamma_A$ |
| $Kr\_m$ | Recovery rate mild symptomatic | 1 / $\gamma_{Sym}$ |
| $Kr\_h$ | Recovery rate hospitalized | 1 / $\gamma_H$ |
| $Kr\_c$ | Recovery rate critical | 1 / $\gamma_C$ |

**S1 Table 2.** Model parameters

| Parameter | Description | Value | Reference |
|---|---|---|---|
| $f_S$ | Fraction symptomatic | 0.5 - 0.7 | [8] |
| $f_{Sm}$ | Fraction mild symptomatic | / | $(1-f_{Ss})$ |
| $f_{Ss}$ | Fraction severe symptomatic | 0.02 - 0.1 | [9] |
| $d_{Sm_t}$ | Fraction of mild symptomatic infections detected (time varying) | 0-0.01 | Calculated from Line List data as described in [10] |
| $d_{Ss_t}$ | Fraction of severe symptomatic infections detected (time varying) | 0.2 - 0.5 | Calculated from Line List data as described in [10] |
| $f_{H_t}$ | Fraction hospitalized | / | $1-f_{C_t}-f_{D_t}$ |
| $f_{C_t}$ | Fraction critical (time varying) | 0.2 - 0.35 | [11] |
| $f_{D_t}$ | Fraction dead (time varying) | / | $\frac{\mathrm{cfr}_t}{f_{Ss}}$ |
| $\mathrm{cfr}_t$ | Case fatality rate (time varying) | 0.01 - 0.04 | [12] |
| $t_{AsP}$ | Time to infectious | 2.4 - 3.5 | [1] |
| $t_P$ | Time exposed to pre-symptomatic | 2.4 - 3.5 | [1] |
| $t_{Sm}$ | Time pre-symptomatic to mild symptoms | 2.4 - 3.5 | [1] [2] |
| $t_{Ss}$ | Time pre-symptomatic to severe symptoms | 3.0 - 4.5 | [1] [2] |
| $t_{Smdet}$ | Time to detection for mild symptomatic | 7 | assumed |
| $t_{Ssdet}$ | Time to detection for severe symptomatic | 2 | assumed |
| $t_H$ | Time to hospitalization | (3, 6) | [3] |
| $t_C$ | Time to critical | 4 - 6 | [4] |
| $t_D$ | Time to deaths | 4 - 6 | [6] |
| $\gamma_A$ | Recovery rate of asymptomatic infections | / | Assumed to be the same as $\gamma_{Sm}$ |
| $\gamma_{Sm}$ | Recovery rate mild symptomatic infections | 7 - 10 | [5] |
| $\gamma_H$ | Recovery rate of hospitalized cases | 4 - 6 | [3] |
| $\gamma_C$ | Recovery rate of critical cases | 8 - 10 | [5] |
| $\delta$ | Reduced infectiousness when detected | 0.1-0.3 | assumed |

**S1 Table 3.** Model equations

| | |
|---|---|
| Transmission and latent period | $\frac{dS}{dt} = -\frac{KiS\left((A + P + Sm_u + Ss_u + H_u + C_u) + (Sm_d + Ss_d)\delta\right)}{N}$ <br> $\frac{dE}{dt} = \frac{KiS\left((A + P + Sm_u + Ss_u + H_u + C_u) + (Sm_d + Ss_d)\delta\right)}{N} - KlE - KsE$ |
| Asymptomatic | $\frac{dA}{dt} = KlE - \gamma_A A$ |
| Pre-symptomatic | $\frac{dP}{dt} = KsE - KsymP - KsysP$ |
| Symptomatic, mild | $\frac{dSm}{dt} = KsymP - \frac{Sm}{t_{Smdet}}$ <br> $\frac{dSm_u}{dt} = \frac{\left(1 - d_{Sm_t}\right)Sm}{t_{Smdet}} - \gamma_{Sym}Sm_u$ <br> $\frac{dSym_d}{dt} = \frac{d_{Sm_t}Sm}{t_{Smdet}} - \gamma_{Sym}Sm_d$ |
| Symptomatic, severe | $\frac{dSs}{dt} = KsysP - \frac{Ss}{t_{Ssdet}}$ <br> $\frac{dSs_u}{dt} = \frac{\left(1 - d_{Ss_t}\right)Ss}{t_{Ssdet}} - Kh1Ss_u - Kh2Ss_u - Kh3Ss_u$ <br> $\frac{dSs_d}{dt} = \frac{d_{Ss_t}Ss}{t_{Ssdet}} - Kh1Ss_d - Kh2Ss_d - Kh3Ss_d$ |
| Hospitalized | $\frac{dH1_d}{dt} = Kh1Ss_d - \gamma_H H1_d$ <br> $\frac{dH2_d}{dt} = Kh2Ss_d - \gamma_H H2_d - KcH2_d$ <br> $\frac{dH3_d}{dt} = Kh3Ss_d - \gamma_H H3_d - KcH3_d$ <br> $\frac{dH1_u}{dt} = Kh1Ss_u - \gamma_H H1_u$ <br> $\frac{dH2_u}{dt} = Kh2Ss_u - \gamma_H H2_u - KcH2_u$ <br> $\frac{dH3_u}{dt} = Kh3Ss_u - \gamma_H H3_u - KcH3_u$ |
| Critical | $\frac{dC2_d}{dt} = KcH2_d - \gamma_C C2_d$ <br> $\frac{dC3_d}{dt} = KcH3_d - KmC3_d$ <br> $\frac{dC2_u}{dt} = KcH2_u - \gamma_C C2_u$ <br> $\frac{dC3_u}{dt} = KcH3_u - KmC3_u$ |
| Deaths | $\frac{dD}{dt} = KmC3_d + KmC3_u$ |
| Recoveries | $\frac{dR}{dt} = \gamma_A A + \gamma_{Sm}Sm_d + \gamma_{Sym}Sm_u + \gamma_H H1_d + \gamma_H H1_u + \gamma_C C2_d + \gamma_C C2_u$ |

### Model parameterization

Data was obtained from Illinois Department of Health (IDPH), with data from individual line list data of COVID-19 cases, Emergency medical resource (EMResource) [13] data for hospital indicators (S1 Table 4).

**S1 Table 4.** Data sources

| Indicator | Use | Source | Ref |
|---|---|---|---|
| Daily deaths | Fitting | I-NEDSS | [14], obtained from IDPH |
| Daily hospitalizations | Fitting | I-NEDSS | [14], obtained from IDPH |
| ICU census | Fitting, capacity limits | EM resource | [13], obtained from IDPH |
| Hospital census | Fitting | EM resource | [13], obtained from IDPH |
| Population | Model input | US Census | [15] |

*Detection rate of mild and severe symptomatic*

The estimated fraction of mild symptomatic detected was obtained from specimen collection data included in line list data shared by IDPH and analyzed on the 28 of July 2020. The detection rates were estimated based on the method described in [10] using an IFR based on Levin et al [16]. Based on the stepwise increase in detection six dates were selected to reflect these in the model.

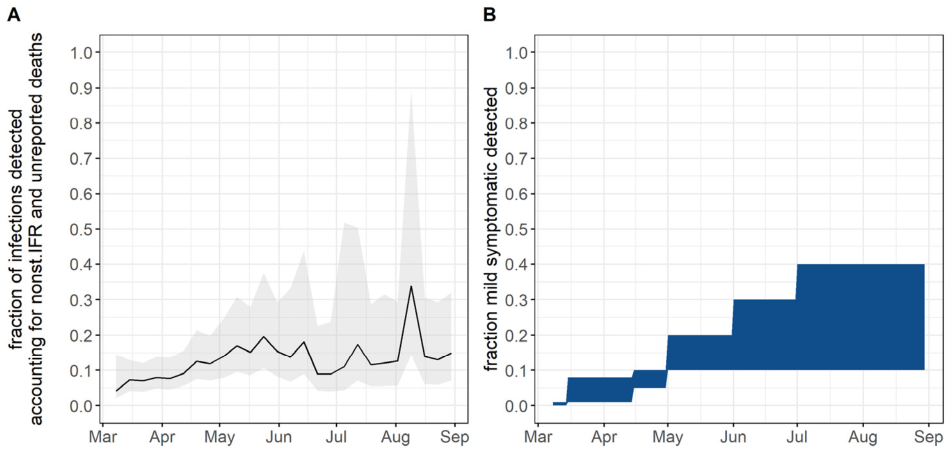

**S1 Fig 2. Detection of mild symptomatic over time in 2020.**
**A)** Estimated fraction of detected infections over time in 2020. **B)** Time-varying model parameter informed by the corresponding trend in data shown in A.

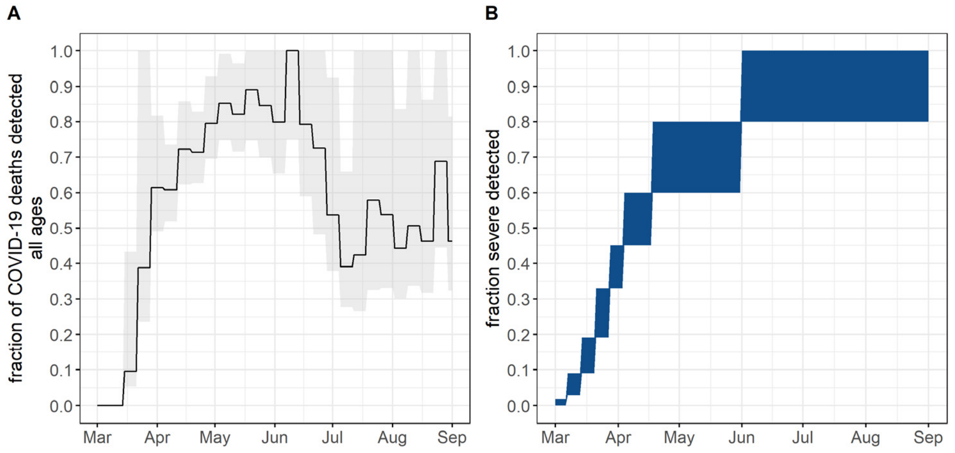

**S1 Fig 3. Detection of severe symptomatic over time in 2020.**

*Fraction critical*

Line list data provided by IDPH, with 21,789 admissions as of June 29, 2020, were analyzed to obtain the fraction critical out of all hospital admissions. The time trend showed a continued decline from 0.75 to <0.1 in July. In the model we used an initial fraction critical rate from literature (see parameter table) and added three universal multipliers in April (*fcritical*=0.13-0.23), May 2020 (*fcritical* = 0.07-0.12) and June 2020 (*fcritical* =0.05-0.09) to reflect the data trend.

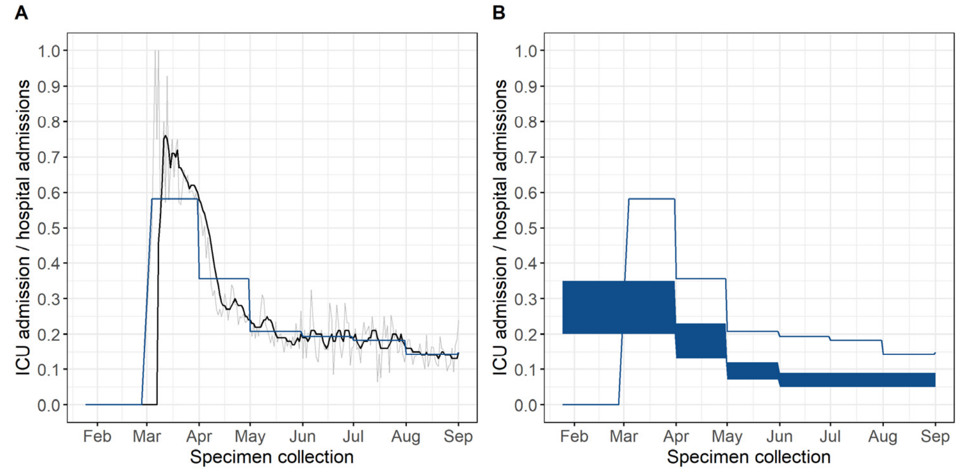

**S1 Fig 4. Fraction critical of admitted cases over time in Illinois over time in 2020.**

**A)** Trends in COVID-19 ICU admissions out of all COVID-19 hospital admissions over time showing raw data per day in grey, 7 day moving average in black and averaged per month in blue. **B)** Averaged monthly trend in COVID-19 ICU admission out of all COVID-19 hospital admissions from line list data compared against model parameters, in which the initial range was informed by literature [11], and the decrease over time by the Illinois specific data. Data source. Individual Line List data from IDPH. Note: not adjusted for delay between specimen collection and hospital admission.

*Case and hospital fatality rate*

The initial case fatality rate was informed from literature [12] and the time-trend was assumed to be relative to the observed trend in the hospital fatality rate calculated from individual line list data. The observed trend in hospital fatality rate suggested a strong decline, dropping by around one third between May and June and another third between June and July. To account for the drop, two scaling factors were added in June and July.

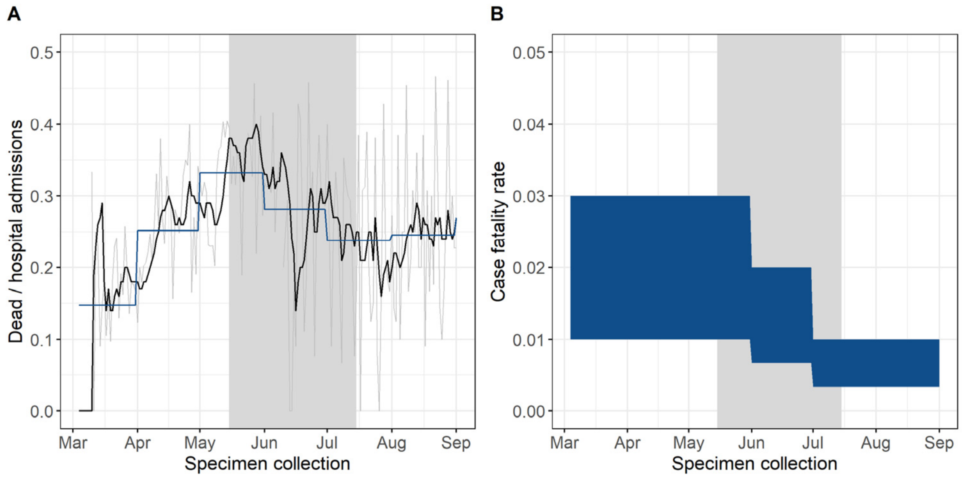

**S1 Fig 5. Hospital admission fatality rate over time in 2020.**
**A)** Trends in COVID-19 deaths out of all COVID-19 hospital admissions over time showing raw data per day in grey, 7 day moving average in black and averaged per month in blue. **B)** Averaged monthly trend in COVID-19 case fatality rate used as model parameter, the initial range was informed by literature [12], and the decrease over time by the Illinois specific data as shown in A. Data source. I-NEDSS.

### Model calibration

In the simulations, the local epidemic was initialized with 10 imported infections and sampled importation dates between January 12 and February 28, 2020. The initial transmission rate was sampled from a range between zero and two on a log-scale. The reduction in transmission after the March 21, 2020, stay-at-home order was sampled from a uniform distribution between 0 and 0.5. After the first simulation that was used to fit the initial epidemic parameters, we ran sequential simulation for every two months with varying transmission rate parameters and time of change to fit monthly mitigation effect sizes. The negative log likelihood was calculated for each simulation trajectory using the *scipy.stats* module in python [17]. Best fit parameter combinations were those that minimized the negative log likelihood of the simulated trajectories, based on a Poisson distribution. In the fitting ICU census and med/surg census, were weighted equally. The model fit was validated against COVID-19-like illness (CLI) hospital admissions data and deaths from IDPH. The final set of simulations ran with the fitted parameters as well as parameter values sampled from uniform distributions for the other parameters and introduced a selection of the top 25% sample parameter combinations that best fit the ICU census data.

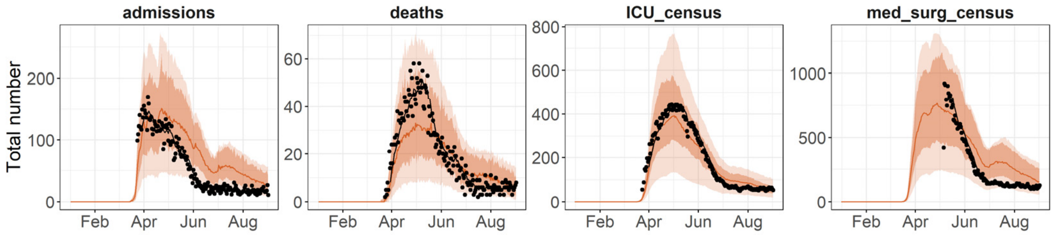

**S1 Fig 6. Comparison to data for four key indicators over time in 2020.**
**A)** Daily hospital admissions from Line List data**. B)** Daily new detected deaths from Line List data. **C)** Daily ICU census from EMResource. **D)** Med/surg census from EMResource**.** The solid line shows the median of the trajectories and the dark shaded area the 90% prediction interval for top 100 trajectories, selected based on fit to ICU census, and the light shaded area the 90% prediction interval for all trajectories.

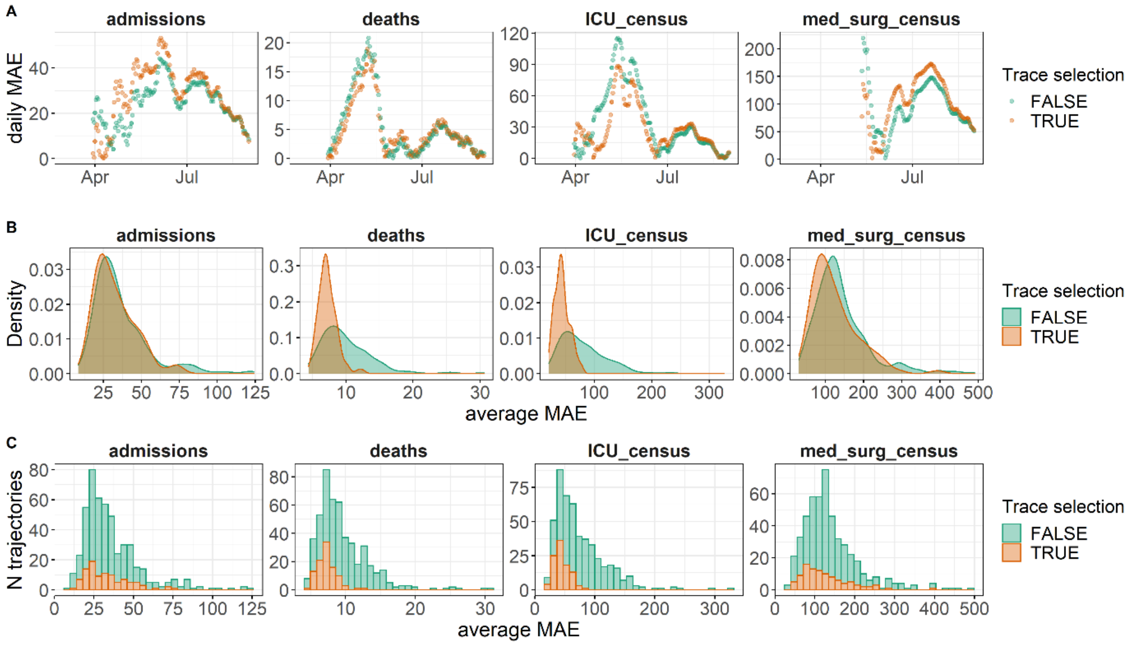

**S1 Fig 7. Selection of best fitting trajectories and change in fitting accuracy per COVID-19 indicator.**
**A)** Daily MAE for the average of the predictions over time between March to August 2020 compared by selection of trajectories for confirmed COVID-19 ICU census, COVID-19 med/surg census and deaths (panels left to right). **B)** Density plot of averaged MAE across the fitting period (March to August 2020) for single trajectories compared by selection of trajectories. **C)** Histogram of average MAE on the x axis for single trajectories compared by selection of trajectories. The 'trace selection' legend and colors indicate the selection step of trajectories, either including all 400 samples (green color) or including the top 100 trajectories that best fit to ICU census (orange color).

### Additional simulation results

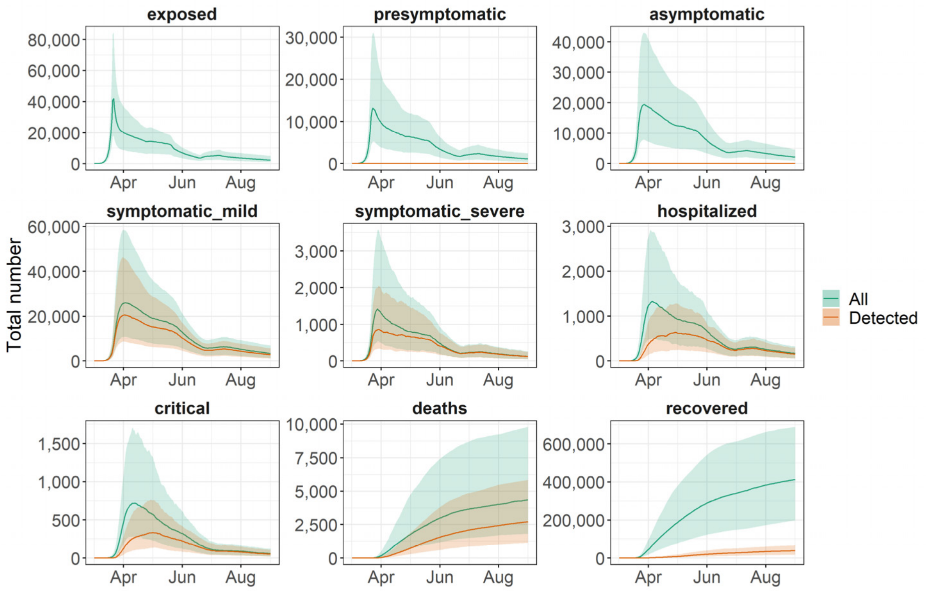

**S1 Fig 8. Daily population count within model compartments predicted for 2020 for selected outcomes showing total population and COVID-19 detections per compartment**.

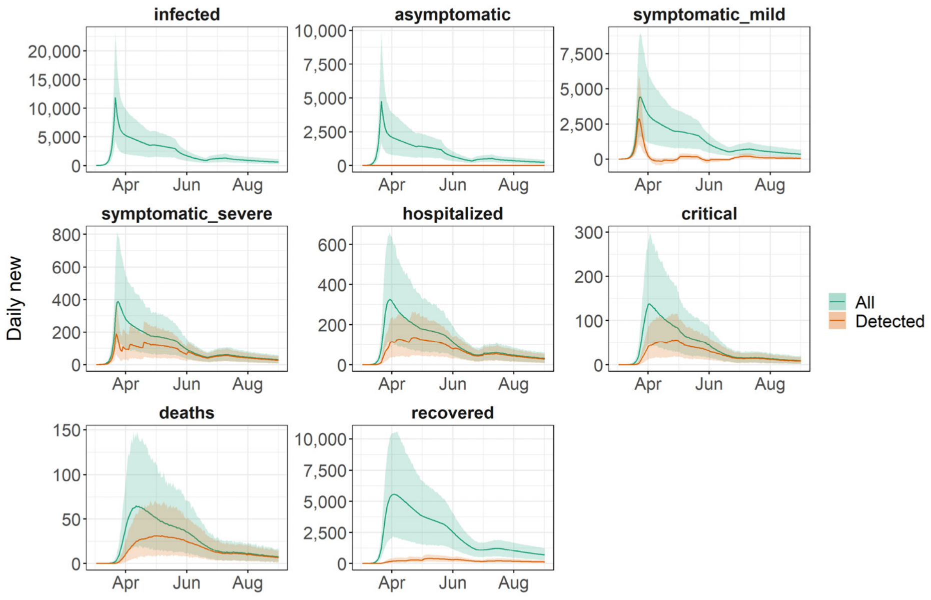

**S1 Fig 9. Daily new model predicted outcomes in 2020 showing total population and COVID-19 detections per compartment**.

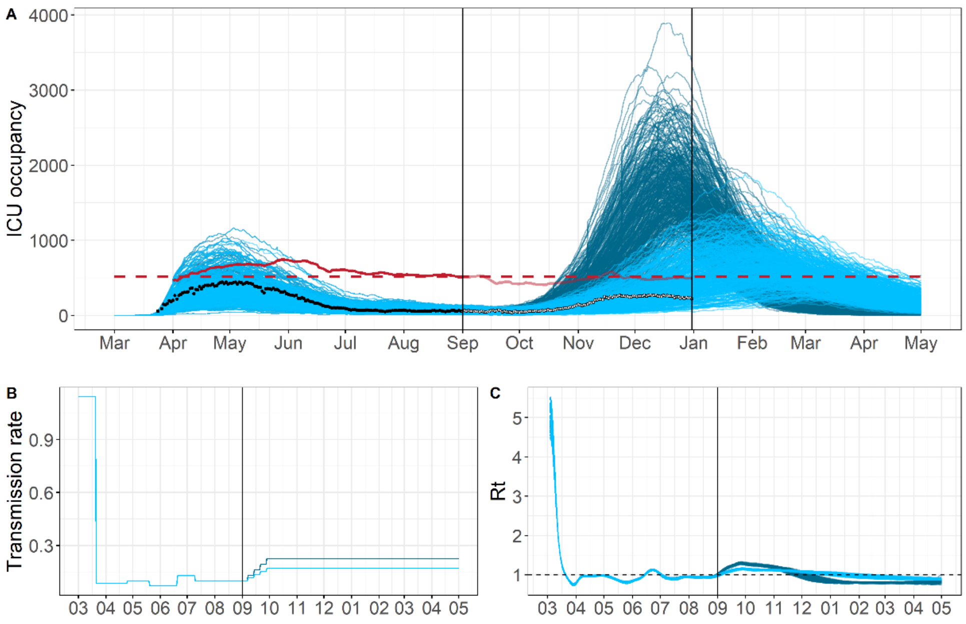

**S1 Fig 10. ICU occupancy trend over time and model fit March 2020 to May 2021.**

**A)** Predicted and observed ICU occupancy for Chicago for 2020. The red line shows the ICU bed capacity for COVID-19 patients (7 day rolling average), assumed to stay constant after September 2020. The two shades of blue indicate a low versus a high level of transmission increase. **B)** Transmission rate parameter fitted to the data before September 2020, and gradually increased afterwards to achieve desired ICU occupancy levels with 100% or 50% exceeding capacity by end of December. **C)** Estimated Rt based on simulated new infections using *epyestim* [18].

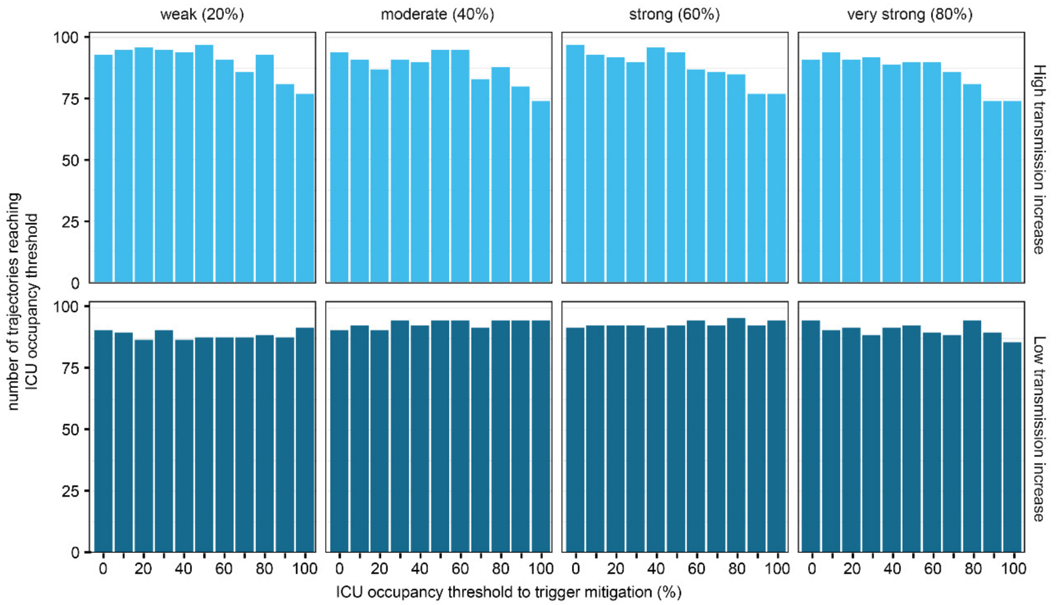

**S1 Fig 11. Number of simulated trajectories that reach ICU occupancy threshold by scenario.**
Showing 100 best fitting trajectories per scenario and ICU occupancy threshold.

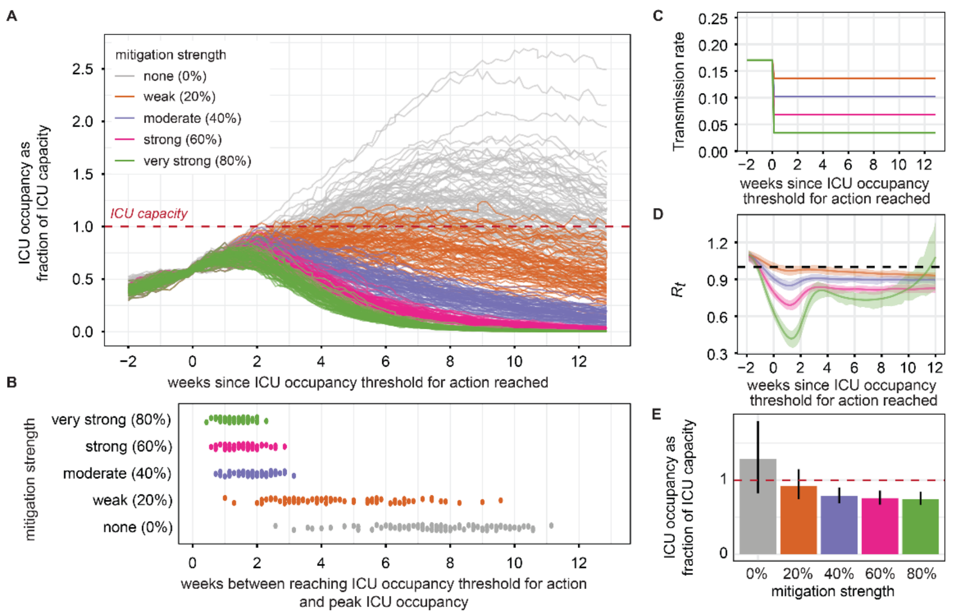

**S1 Fig 12. Projected outcomes under no mitigation compared to four mitigation strengths.**
**A)** Projected ICU census over time relative to the time when ICU occupancy reached the ICU occupancy threshold. **B)** Timing of peak ICU occupancy relative to the time when ICU occupancy reached the ICU occupancy threshold. **C)** Reduction in transmission rate due to immediate mitigation after reaching a 60% occupancy threshold. **D)** Estimated mean Rt with 90%PI uncertainty intervals. **E)** Peak ICU occupancy, with mean and 90% PI error bars, by mitigation strength. All mitigations were implemented 1 day after reaching a 60% occupancy threshold.

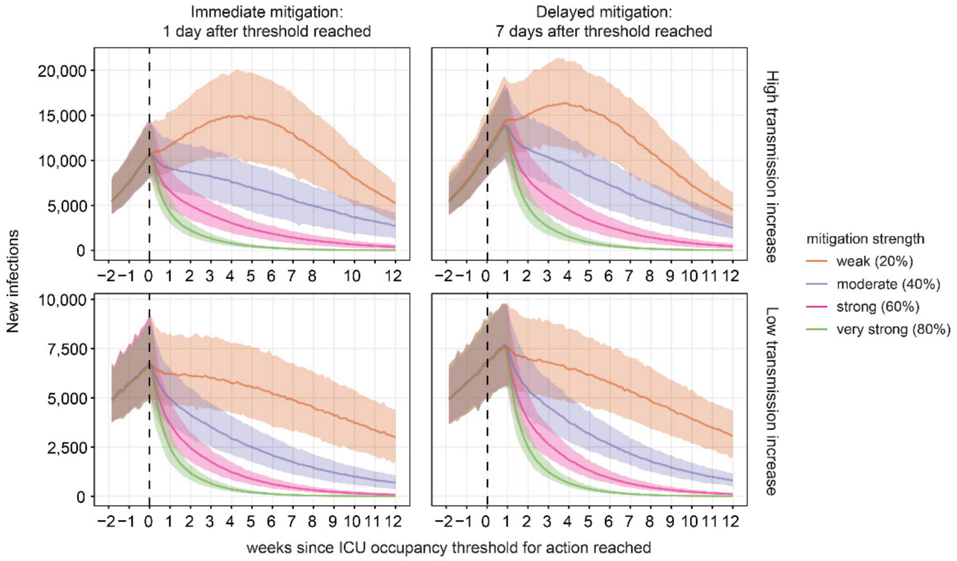

**S1 Fig 13. Projected daily new infections by scenario.**
Mitigation is triggered when reaching 60% ICU occupancy (dashed vertical line).

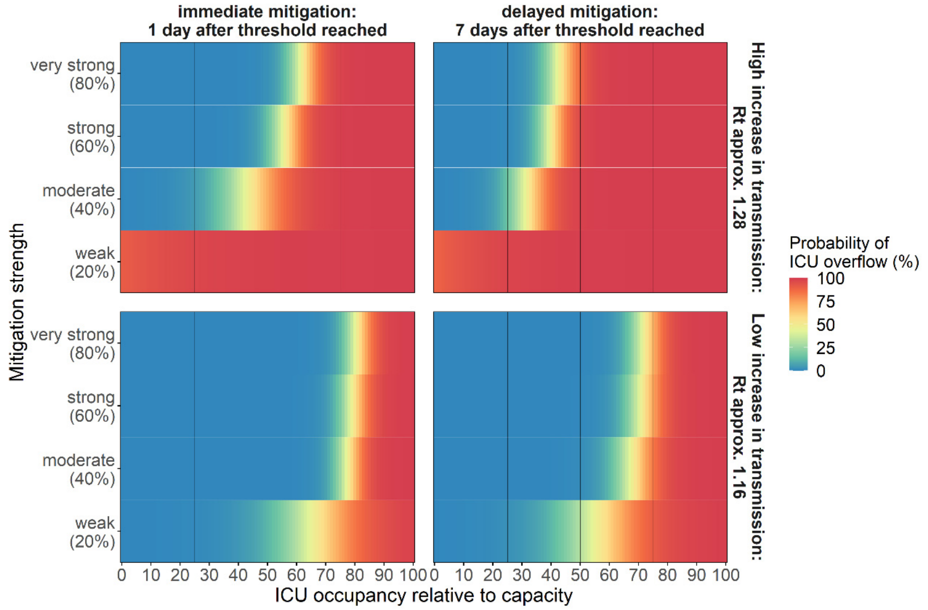

**S1 Fig 14. Probability of exceeding ICU capacity by ICU occupancy threshold, mitigation strength and timing**. The fill color shows the probability of exceeding ICU capacity with low probability in blue and high probability in red as indicated by the numbers within each tile. A logistic regression was used to predict probability for occupancy thresholds between the simulated intervals. The black vertical lines indicate an ICU occupancy threshold of 25, 50 and 75%.

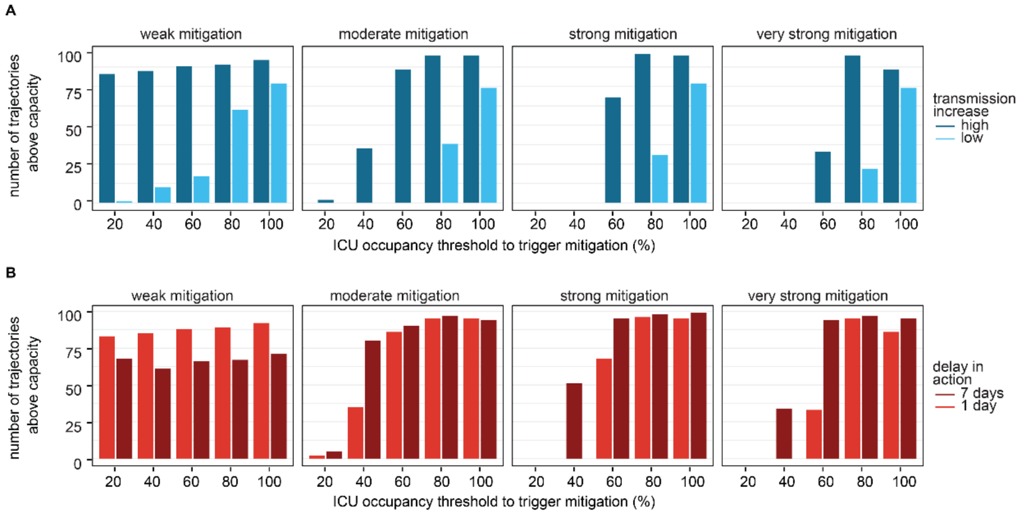

**S1 Fig 15. Number of trajectories above capacity** for different scenarios by ICU occupancy threshold.
**A)** Number of trajectories above capacity per ICU occupancy threshold triggering immediate mitigation by transmission increase level. **B)** Number of trajectories above capacity per ICU occupancy threshold triggering immediate mitigation by delay at higher transmission increase. Total N=100 best fitting trajectories.

### Hospital data trends in Chicago and post-analysis predictions

*ICU occupancies*

The hospital census data provided information on hospital and ICU bed occupancies categorized by COVID-19 disease status (yes/no). All ICU beds that exist is referred to as total ICU capacity. Most of these beds are occupied by patients with non-COVID-19 related diseases, leaving a subset of the total capacity available for COVID-19 patients, which is referred to as ICU capacity in the analysis. When subtracting the number of beds occupied by COVID-19 patients from the ICU capacity (for COVID-19 patients) the output is the number of beds available for new patients. The figure below shows the trends in all of these ICU indicators over time, with a marked scale up of beds early during the pandemic, constant total capacity during the summer at higher numbers than before the pandemic and some scale down and up during fall as COVID-19 ICU occupancy started to increase again (S1 Fig 16).

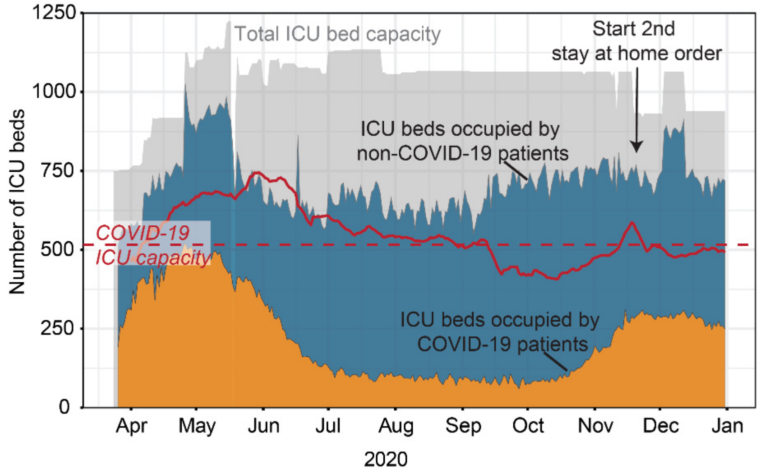

**S1 Fig 16. Total ICU capacity, availability and occupancy for all and COVID-19 patients over time in 2020.** The arrow points at Nov 20, 2020, the start of the second stay at home order [19]. The orange area shows number of ICU beds occupied by COVID-19 patients, the blue area additional beds occupied by non-COVID-19 patients and the grey area the number of unoccupied (available beds). The ICU capacity for COVID-19 patients is calculated by subtracting the ICU occupancy of non-COVID-19 patients from the total ICU capacity.

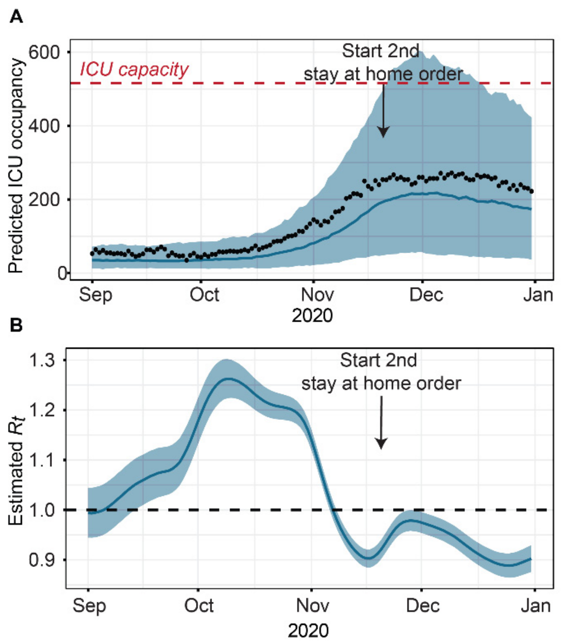

**S1 Fig 17. ICU occupancy and estimated Rt using model calibrated until December 2020.**
**A)** Predicted ICU occupancy in blue and reported ICU census in black. The shaded area shows the 95%PI. **B)** Estimated Rt based on predicted new infections showing median and 95%CI based on uncertainty in estimation method (*epyestim*).

*Home location of hospital patients*

Weekly hospital admission data from the Northwestern Memorial Hospital in Chicago between March to November 2020 was analyzed to obtain estimates on fraction of patients from Chicago or surrounding areas. The trend in total number of admissions shows a similar trend to the ICU census shown in S1 Fig 18, with a step increase between March to May, constantly fewer admissions between June to October, before a new step increase in admissions (S1 Fig 18A). Overall, between around 70 to 80% patients were from Chicago areas, with the remaining 20-30% from the surrounding Cook County or other regions in Illinois and very few from outside the State (S1 Fig 18B).

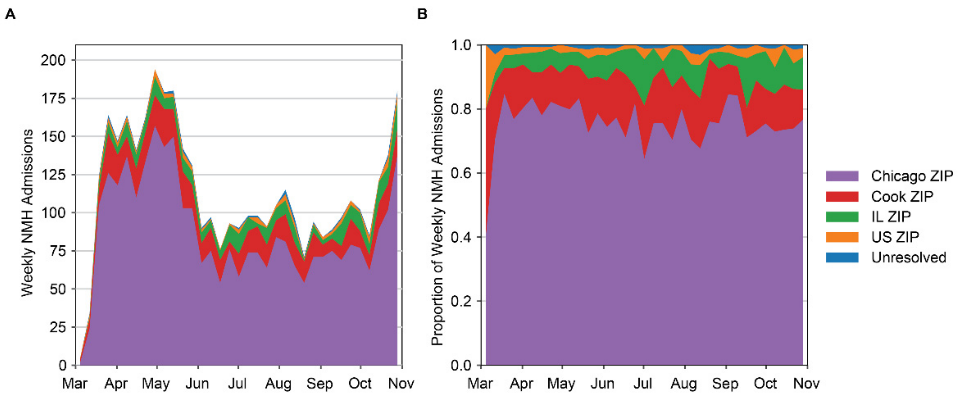

**S1 Fig 18. Northwestern Memorial Hospital patients by ZIP code in Chicago and surrounding areas in 2020.**
**A)** Weekly hospital admissions. **B)** Proportion of weekly hospital admissions by ZIP code.

### Comparison of simulated reproductive numbers and mitigation effect to other studies

*Basic reproductive number and early mitigation effects*

Overall, our model estimated a relatively high basic reproductive number of $R_0$ = 5 (90% PI 4.57-5.41) when comparing to other model estimates for Illinois [20,21] or early reproductive numbers estimated from data in China [22–25], that lie between 2-3. However, basic reproductive numbers in Chicago are likely higher as the rest of Illinois due to population density [26] and basic reproductive numbers of 4-6 or higher have been estimated in other states in the US [21] and are more similar to estimates for New York [20]. It might also be that the basic reproductive number was overestimated in our model, as we estimated the infection importation date, the initial transmission rate, as well as mitigation effect size within the same model calibration for February-March 2020, and multiple combinations exists that can reproduce observed trends in the data. The initial mitigation effect size of 92% is relatively high compared to 62% estimated in a pooled analysis [27]. Obtaining accurate estimates on the effect size of mitigation is challenging as mitigation effects are highly heterogeneous across settings [28].

*Simulated transmission increase in fall 2020*

In our simulation, the transmission increase levels represented a wide range of epidemic growth scenarios to demonstrate robustness of the trigger thresholds that account for the large uncertainty in the transmission dynamics. Studies suggested that COVID-19 transmission was likely to increase during fall and winter months, due to a mix of environmental, behavioral, and political factors [3–5]. The increase in Oct-Nov 2020 was observed in many States across the US [29].

*Simulated reduction in effective reproduction number due to mitigation*

In our analysis we selected a 20, 40, 60 and 80% reduction in the transmission rate to obtain predictions for a wide range of possible mitigation effect sizes. The corresponding estimated reduction in the effective reproductive numbers similar between both transmission increase scenarios but lower compared to the reduction in the transmission rate (S1 Table 5, S1 Fig 19). Another modeling study conducted for regions in the UK estimated mitigation impact between 2% to 44% reduction in Rt, depending on mitigation tier, and their post-hoc analysis estimated a reduction of 22% [30].

**S1 Table 5.** Comparison of simulated reproductive numbers and mitigation effect to other studies*

| | Simulation | | Other studies | |
|---|---|---|---|---|
| | **Low transmission increase scenario** | **High transmission increase scenario** | **Estimate** | **Reference** |
| $R_0$ | 5.0 (4.57-5.41) | 5.0 (4.57-5.41) | ~2.5 (~2-4) | [20] (Illinois) |
| | | | ~ 3.09 (2.86, 3.3) | [21] (Illinois) |
| | | | ~4.7 (3.8-5.5) | [20] (New York) |
| | | | 1.66 (1.35-2.11) | [26] (USA, 310 counties) |
| | | | 4.08 (3.09–5.39) | [27] (Pooled, global estimate) |

| | | | | |
|---|---|---|---|---|
| % Decrease in $R_t$ due to mitigation during first wave | 92.5% (from ~5 to 0.74) | 92.5% (from ~5 to 0.74) | 30-45% (1.4) | [31] (Washington, USA) |
| | | | 29%-64% (NA) | [32] (UK) |
| | | | 62% (from 4.08 to 1.51) | [27] (Pooled, global estimate) |
| | | | 74% (0.62, 0.37–0.89) | [33] (UK) |
| | | | 85% (NA) | [5] (Shenzhen China) |
| | | | 92% (from 3.82 to 0.3) | [34] (Wuhan, China) |
| | | | 95% (from 10.3 to 0.44) | [35] (Texas, USA) |
| % Increase in $R_t$ during second wave | 14% (1.14, 1.12 - 1.16) | 28% (1.28, 1.25- 1.30) | / | / |
| % Decrease in $R_t$ due to mitigation during second wave (new $R_t$ estimate under mitigation) | 12%, 23%, 37%, 63% (from ~1.09 to 0.96, 0.84, 0.67, 0.40) ** | 16%, 25%, 39%, 62% (from ~1.23 to 1.03, 0.92, 0.75, 0.46) ** | 2-44% (NA) Range across tiers | [30] (England) |
| | | | 22% (1.0, 0.6-1.2) | [30] (England) Post-hoc estimation |

*) crude, non-systematic, comparison of magnitude in estimates across different estimation methods, geographical locations, and resolution as well as calendar times.
**) for 20%, 40%, 60%, 80% reduction in transmission rate triggered at 60% of ICU occupancy.

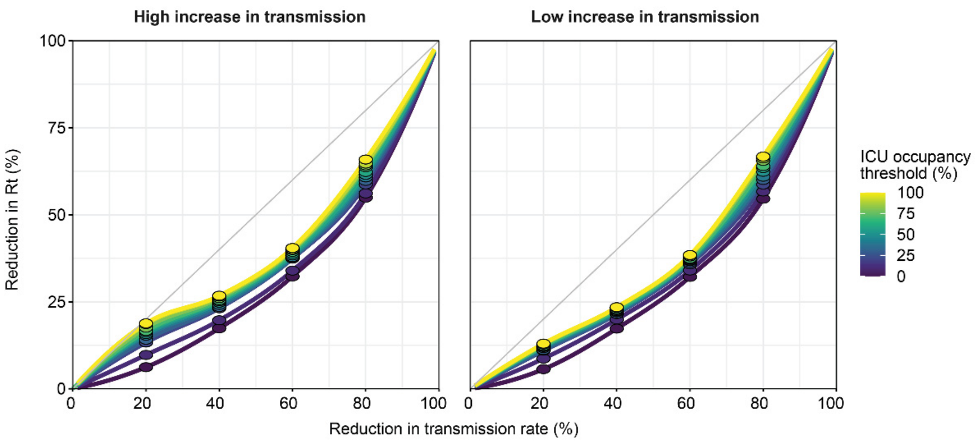

**S1 Fig 19. Estimated relationship between reduction in transmission rate and in $R_t$.**
The points show the reduction in $R_t$ after two weeks after mitigation was triggered immediately after reaching the ICU occupancy threshold, averaged across simulated trajectories. The lines show fitted loess regression after adding constrained to go through 0 and 100%. $R_t$ was estimated using projected total daily new infections and the python module *epyestim* [18]. The $R_t$ estimates 2 weeks prior to mitigation ranged from 1.08 to 1.40 for the higher level of transmission increase scenario and from 0.94 to 1.19 for the lower level of transmission increase.

### Simulation completeness and selection of trajectories

**S1 Table 6.** Number of simulated trajectories per simulation scenario (N = 4400, 400 samples * 11 ICU occupancy thresholds)

| **Simulation scenario** | | | **N trajectories** | | | | | | **% trajectories** | | | | | |
|---|---|---|---|---|---|---|---|---|---|---|---|---|---|---|
| | | | all trajectories | | | best fitting trajectories | | | all trajectories | | | best fitting trajectories | | |
| **reopen** | **delay** | **mitigation** | success | reached ICU occupancy threshold* | reached peak in ICU occupancy* | top traces | reached ICU occupancy threshold | reached peak in ICU occupancy | success | reached ICU occupancy threshold | reached peak in ICU occupancy | top traces | reached ICU occupancy threshold | reached peak in ICU occupancy |
| High | 1 day | weak | 4007 | 4007 | 4007 | 979 | 979 | 979 | 0.911 | 1 | 1 | 0.244 | 1 | 1 |
| High | 1 day | moderate | 4114 | 4114 | 4114 | 1030 | 1030 | 1030 | 0.935 | 1 | 1 | 0.25 | 1 | 1 |
| High | 1 day | strong | 3998 | 3998 | 3998 | 1028 | 1028 | 1028 | 0.909 | 1 | 1 | 0.257 | 1 | 1 |
| High | 1 day | very strong | 4124 | 4124 | 4124 | 1002 | 1002 | 1002 | 0.937 | 1 | 1 | 0.243 | 1 | 1 |
| High | 7 days | weak | 4164 | 2909 | 2909 | 726 | 726 | 726 | 0.661 | 1 | 1 | 0.25 | 1 | 1 |
| High | 7 days | moderate | 3982 | 4131 | 4131 | 1027 | 1027 | 1027 | 0.939 | 1 | 1 | 0.249 | 1 | 1 |
| High | 7 days | strong | 4130 | 4236 | 4236 | 1059 | 1059 | 1059 | 0.963 | 1 | 1 | 0.25 | 1 | 1 |
| High | 7 days | very strong | 3995 | 4286 | 4286 | 1071 | 1071 | 1071 | 0.974 | 1 | 1 | 0.25 | 1 | 1 |
| Low | 1 day | weak | 2909 | 3937 | 4164 | 1044 | 996 | 1044 | 0.946 | 0.945 | 1 | 0.251 | 0.954 | 1 |
| Low | 1 day | moderate | 4131 | 3760 | 3982 | 1014 | 966 | 1014 | 0.905 | 0.944 | 1 | 0.255 | 0.953 | 1 |
| Low | 1 day | strong | 4236 | 3892 | 4130 | 1024 | 972 | 1024 | 0.939 | 0.942 | 1 | 0.248 | 0.949 | 1 |
| Low | 1 day | very strong | 4286 | 3784 | 3995 | 995 | 950 | 995 | 0.908 | 0.947 | 1 | 0.249 | 0.955 | 1 |
| Low | 7 days | weak | 4193 | 3955 | 4193 | 1060 | 1006 | 1060 | 0.953 | 0.943 | 1 | 0.253 | 0.949 | 1 |
| Low | 7 days | moderate | 3923 | 3705 | 3923 | 961 | 911 | 961 | 0.892 | 0.944 | 1 | 0.245 | 0.948 | 1 |
| Low | 7 days | strong | 4352 | 4112 | 4352 | 1090 | 1036 | 1090 | 0.989 | 0.945 | 1 | 0.25 | 0.95 | 1 |
| Low | 7 days | very strong | 4114 | 3898 | 4114 | 1026 | 977 | 1026 | 0.935 | 0.947 | 1 | 0.249 | 0.952 | 1 |

* ) Reached ICU occupancy threshold and peak by May 1, 2021.

**S1 Table 7.** Projected peak in ICU occupancy and time events (median and 90% PI)

| Transmission increase | Mitigation | delay | ICU threshold | ICU occupancy at peak (relative to capacity) | | | Time Oct 1 to ICU peak (days) | | | Time trigger to peak (days) | | | Time Oct 1 to trigger (days) | | |
|---|---|---|---|---|---|---|---|---|---|---|---|---|---|---|---|
| | | | | median | lower | upper | median | lower | upper | median | lower | upper | median | lower | upper |
| High | weak | 1 day | 0.4 | 1.1 | 2.0 | 3.2 | 103 | 118 | 136 | 47 | 61 | 75 | 46 | 57 | 76 |
| High | weak | 1 day | 0.6 | 1.3 | 2.0 | 3.1 | 97 | 115 | 131 | 34 | 48 | 64 | 54 | 65 | 85 |
| High | weak | 1 day | 0.8 | 1.4 | 2.2 | 3.2 | 97 | 110 | 126 | 26 | 42 | 52 | 59 | 69 | 86 |
| High | weak | 1 day | 1 | 1.5 | 2.3 | 3.2 | 96 | 108 | 130 | 20 | 33 | 45 | 62 | 75 | 106 |
| High | moderate | 1 day | 0.4 | 0.7 | 0.9 | 1.3 | 80 | 93 | 113 | 19 | 33 | 62 | 46 | 57 | 75 |
| High | moderate | 1 day | 0.6 | 0.9 | 1.2 | 1.6 | 79 | 92 | 105 | 14 | 26 | 44 | 54 | 64 | 88 |
| High | moderate | 1 day | 0.8 | 1.2 | 1.5 | 1.9 | 79 | 93 | 105 | 14 | 21 | 35 | 59 | 70 | 86 |
| High | moderate | 1 day | 1 | 1.4 | 1.7 | 2.2 | 81 | 94 | 120 | 12 | 20 | 27 | 62 | 75 | 106 |
| High | strong | 1 day | 0.4 | 0.6 | 0.8 | 0.9 | 61 | 72 | 90 | 11 | 15 | 20 | 46 | 57 | 76 |
| High | strong | 1 day | 0.6 | 0.9 | 1.0 | 1.2 | 69 | 80 | 97 | 10 | 15 | 20 | 54 | 64 | 88 |
| High | strong | 1 day | 0.8 | 1.1 | 1.4 | 1.6 | 75 | 83 | 97 | 10 | 14 | 18 | 61 | 69 | 86 |
| High | strong | 1 day | 1 | 1.3 | 1.6 | 1.8 | 77 | 89 | 116 | 10 | 13 | 17 | 62 | 75 | 106 |
| High | very strong | 1 day | 0.4 | 0.6 | 0.7 | 0.8 | 57 | 70 | 87 | 10 | 12 | 16 | 46 | 57 | 75 |
| High | very strong | 1 day | 0.6 | 0.8 | 1.0 | 1.1 | 65 | 76 | 95 | 9 | 12 | 16 | 54 | 65 | 88 |
| High | very strong | 1 day | 0.8 | 1.1 | 1.3 | 1.5 | 71 | 81 | 97 | 9 | 12 | 15 | 59 | 70 | 86 |
| High | very strong | 1 day | 1 | 1.3 | 1.5 | 1.7 | 74 | 85 | 102 | 9 | 11 | 14 | 62 | 75 | 93 |
| High | weak | 7 days | 0.4 | 1.3 | 2.0 | 3.2 | 102 | 114 | 127 | 41 | 56 | 67 | 48 | 59 | 75 |
| High | weak | 7 days | 0.6 | 1.4 | 2.2 | 3.1 | 94 | 110 | 123 | 27 | 45.5 | 53 | 53 | 66 | 85 |
| High | weak | 7 days | 0.8 | 1.6 | 2.4 | 3.6 | 95 | 110 | 124 | 25 | 40 | 47 | 60 | 70 | 87 |
| High | weak | 7 days | 1 | 1.6 | 2.5 | 3.5 | 94 | 107 | 132 | 21 | 32 | 42 | 62 | 75 | 111 |
| High | moderate | 7 days | 0.4 | 0.9 | 1.2 | 1.6 | 80 | 91 | 105 | 22 | 32 | 50 | 46 | 58 | 76 |
| High | moderate | 7 days | 0.6 | 1.2 | 1.6 | 2.0 | 80 | 92 | 108 | 19 | 26 | 37 | 53 | 65 | 85 |
| High | moderate | 7 days | 0.8 | 1.4 | 1.9 | 2.5 | 84 | 94 | 109 | 19 | 25 | 32 | 59 | 69 | 87 |
| High | moderate | 7 days | 1 | 1.5 | 2.2 | 2.7 | 88 | 96 | 120 | 16 | 23 | 29 | 62 | 74 | 105 |
| High | strong | 7 days | 0.4 | 0.8 | 1.0 | 1.3 | 65 | 79 | 94 | 16 | 20 | 25 | 46 | 58 | 76 |
| High | strong | 7 days | 0.6 | 1.0 | 1.4 | 1.6 | 73 | 85 | 104 | 16 | 20 | 25 | 53 | 65 | 87 |
| High | strong | 7 days | 0.8 | 1.3 | 1.7 | 2.1 | 79 | 88 | 105 | 16 | 19 | 24 | 59 | 69 | 87 |

| | | | | | | | | | | | | | | | |
|---|---|---|---|---|---|---|---|---|---|---|---|---|---|---|---|
| High | strong | 7 days | 1 | 1.5 | 2.0 | 2.4 | 81 | 93 | 120 | 15 | 18 | 22 | 62 | 74 | 105 |
| High | very strong | 7 days | 0.4 | 0.8 | 0.9 | 1.2 | 64 | 76 | 92 | 16 | 18 | 20 | 47 | 58 | 75 |
| High | very strong | 7 days | 0.6 | 1.0 | 1.3 | 1.6 | 70 | 83 | 104 | 14 | 17 | 21 | 53 | 65 | 87 |
| High | very strong | 7 days | 0.8 | 1.3 | 1.6 | 2.0 | 77 | 87 | 102 | 15 | 17 | 20 | 59 | 69 | 87 |
| High | very strong | 7 days | 1 | 1.5 | 1.9 | 2.3 | 79 | 92 | 120 | 14 | 17 | 20 | 62 | 74 | 105 |
| Low | weak | 1 day | 0.4 | 0.5 | 0.7 | 1.1 | 108 | 130 | 158 | 12 | 40.5 | 76 | 65 | 86 | 130 |
| Low | weak | 1 day | 0.6 | 0.6 | 0.9 | 1.1 | 108 | 132 | 162 | 8 | 26 | 54 | 77 | 101 | 144 |
| Low | weak | 1 day | 0.8 | 0.9 | 1.0 | 1.4 | 111 | 131 | 167 | 5 | 18 | 36 | 91 | 110 | 158 |
| Low | weak | 1 day | 1 | 1.0 | 1.2 | 1.5 | 115 | 136 | 170 | 2 | 16 | 33 | 91 | 119 | 158 |
| Low | moderate | 1 day | 0.4 | 0.4 | 0.5 | 0.7 | 80 | 104 | 158 | 9 | 15 | 24 | 65 | 87 | 145 |
| Low | moderate | 1 day | 0.6 | 0.6 | 0.8 | 0.9 | 89 | 112 | 152 | 5 | 12 | 19 | 75 | 101 | 144 |
| Low | moderate | 1 day | 0.8 | 0.8 | 1.0 | 1.1 | 103 | 123 | 165 | 3 | 11 | 17 | 92 | 110 | 158 |
| Low | moderate | 1 day | 1 | 1.0 | 1.2 | 1.4 | 102 | 130 | 170 | 1 | 10 | 16 | 91 | 119 | 162 |
| Low | strong | 1 day | 0.4 | 0.4 | 0.5 | 0.6 | 74 | 95 | 150 | 6 | 11 | 16 | 64 | 85 | 145 |
| Low | strong | 1 day | 0.6 | 0.6 | 0.7 | 0.9 | 87 | 112 | 149 | 5 | 10 | 15 | 75 | 101 | 144 |
| Low | strong | 1 day | 0.8 | 0.8 | 1.0 | 1.1 | 101 | 122 | 163 | 3 | 10 | 13 | 91 | 112 | 158 |
| Low | strong | 1 day | 1 | 1.0 | 1.1 | 1.3 | 101 | 130 | 174 | 1 | 9 | 13 | 91 | 122 | 164 |
| Low | very strong | 1 day | 0.4 | 0.4 | 0.5 | 0.6 | 74 | 94 | 137 | 4 | 10 | 13 | 65 | 85 | 130 |
| Low | very strong | 1 day | 0.6 | 0.6 | 0.7 | 0.8 | 87 | 112 | 146 | 5 | 9 | 13 | 77 | 101 | 139 |
| Low | very strong | 1 day | 0.8 | 0.8 | 1.0 | 1.1 | 100 | 119 | 162 | 3 | 9 | 11 | 91 | 110 | 156 |
| Low | very strong | 1 day | 1 | 1.0 | 1.1 | 1.3 | 101 | 129 | 168 | 1 | 8 | 11 | 91 | 122 | 162 |
| Low | weak | 7 days | 0.4 | 0.5 | 0.7 | 1.2 | 109 | 131 | 173 | 19 | 43 | 69 | 64 | 86 | 145 |
| Low | weak | 7 days | 0.6 | 0.7 | 0.9 | 1.2 | 107 | 132 | 162 | 13 | 27 | 49 | 76 | 101 | 143 |
| Low | weak | 7 days | 0.8 | 0.9 | 1.1 | 1.5 | 115 | 135 | 168 | 9 | 21 | 37 | 90 | 109 | 158 |
| Low | weak | 7 days | 1 | 1.0 | 1.3 | 1.7 | 116 | 139 | 177 | 5 | 18 | 34 | 91 | 119 | 164 |
| Low | moderate | 7 days | 0.4 | 0.5 | 0.6 | 0.8 | 84 | 106 | 161 | 13 | 19.5 | 26 | 64 | 86 | 144 |
| Low | moderate | 7 days | 0.6 | 0.7 | 0.9 | 1.0 | 97 | 121 | 162 | 10 | 18 | 25 | 77 | 101 | 143 |
| Low | moderate | 7 days | 0.8 | 0.9 | 1.1 | 1.4 | 110 | 127 | 167 | 9 | 16.5 | 22 | 91 | 110 | 158 |
| Low | moderate | 7 days | 1 | 1.0 | 1.3 | 1.5 | 111 | 135 | 175 | 5 | 16 | 21 | 91 | 119 | 161 |
| Low | strong | 7 days | 0.4 | 0.5 | 0.6 | 0.8 | 81 | 102 | 158 | 12 | 17 | 23 | 64 | 86 | 145 |
| Low | strong | 7 days | 0.6 | 0.6 | 0.8 | 1.0 | 94 | 116 | 162 | 8 | 16 | 19 | 76 | 101 | 143 |

| | | | | | | | | | | | | | | | |
|---|---|---|---|---|---|---|---|---|---|---|---|---|---|---|---|
| Low | strong | 7 days | 0.8 | 0.9 | 1.1 | 1.3 | 105 | 124 | 167 | 9 | 15 | 19 | 90 | 110 | 158 |
| Low | strong | 7 days | 1 | 1.0 | 1.2 | 1.5 | 109 | 134 | 173 | 6 | 15 | 18 | 94 | 119 | 161 |
| Low | very strong | 7 days | 0.4 | 0.5 | 0.6 | 0.7 | 78 | 101 | 158 | 11 | 16 | 20 | 64 | 86 | 145 |
| Low | very strong | 7 days | 0.6 | 0.7 | 0.8 | 1.0 | 91 | 114 | 152 | 8 | 15 | 18 | 77 | 101 | 139 |
| Low | very strong | 7 days | 0.8 | 0.9 | 1.1 | 1.2 | 104 | 124 | 167 | 9 | 14 | 17 | 90 | 109 | 157 |
| Low | very strong | 7 days | 1 | 1.0 | 1.2 | 1.4 | 105 | 133 | 173 | 5 | 14 | 17 | 91 | 119 | 161 |
